# Trends in the Utilization of Breast, Cervical, and Colorectal Cancer Screening from 2010 to 2019 Among a Commercially Insured Population Using the MarketScan Commercial Claims Database

**DOI:** 10.64898/2026.08.09.26360037

**Authors:** Jingjing Sun, Ricky Wat, K. Davina Frick, Xiangrong Kong, Hailun Liang, Clifton Chow, Leiyu Shi

## Abstract

**Introduction:** Breast, cervical, and colorectal cancer screening guidelines changed substantially between 2010 and 2019. We examined trends in the annual utilization of these screenings among commercially insured enrollees in the United States from 2010 to 2019 by age group, geographic region, and screening modality.

**Methods:** We conducted a retrospective, serial cross-sectional analysis of the MarketScan Commercial Claims Database from 2010 through 2019, comprising approximately 141.2 million privately insured enrollees. Annual screening rates, defined as the proportion of eligible enrollees receiving a given test within each calendar year, were estimated for cervical, breast, and colorectal cancer using procedure codes, stratified by age group, screening modality, and geographic residence. These reflect annual utilization rather than up-to-date (guideline-concordant) screening. Temporal trends were evaluated using two-sided Poisson regression, and urban-rural disparities in 2019 were assessed using multivariate generalized estimating equations.

**Results:** Cancer screening utilization remained stagnant or declined across all three cancer types over the study period. Among women aged 30-64 years, cervical cytology alone declined substantially from 28.2% in 2010 to 8.8% in 2019, while co-testing increased from 11.4% to 20.3%. Screening mammography among women aged 50-64 showed minimal change, remaining stable at 45.7% in 2010 and 45.8% in 2019. Colorectal cancer screening across enrollees aged <64 decreased modestly from 7.7% in 2010 to 6.5% in 2019, with a more pronounced decline among adults aged 45-49 years. Across all three cancer types, screening utilization was higher among urban residents than rural residents, with incidence rate ratios ranging from 1.02 to 1.05 in 2019.

**Conclusions:** Utilization of cervical, breast, and colorectal cancer screening among commercially insured adults did not improve between 2010 and 2019. Persistent urban-rural disparities highlight ongoing gaps in preventive care delivery. Targeted interventions may help improve screening utilization, particularly in rural and underserved populations.

## Introduction

Cancer screening is an important preventive care service designed to enable early detection and treatment of cancer. Over the past decade, considerable changes have occurred in US guidelines and recommendations for breast, cervical, and colorectal cancer screening, influenced by the introduction of new screening technology and by improved understanding of causal mechanisms behind cancer development. These guideline changes include changing the age to initiate screening, inclusion of new test modalities, or modifying screening interval length considered to be up to date, which may vary based on individual age and risk profile.^1-4^

Guidelines and recommendations for cancer screening vary across professional organizations and have evolved during the study period, creating complexity in implementation.^1,5^ For cervical cancer, major guidelines including the United States Preventive Services Task Force (USPSTF), American Cancer Society (ACS), and American College of Obstetricians and Gynecologists (ACOG) endorsed co-testing in addition to cytology alone for women aged 30-65 by 2012, with primary human papillomavirus (HPV) testing added as an option in 2018.^4,5^ For breast cancer screening, recommendations differ regarding screening initiation age and frequency, stirring debate among healthcare providers and patients about the relative benefits and harms at different age groups.^6^ These variations in recommendations across organizations have created complexity in how guidelines are implemented. It remains unclear how complex changes in different guidelines and recommendations may have impacted cancer screening utilization during this period.^4^

Observing changes in cancer screening patterns over time can offer valuable insight into how guidelines are influencing patient and healthcare provider behaviors. Although large-scale public health survey databases such as the National Health Interview Survey (NHIS), Health Information National Trends Survey (HINTS), and Behavioral Risk Factor Surveillance System (BRFSS) are widely used to monitor changes in breast, cervical, and colorectal screening at the national and state level, there are limitations to these survey databases, including reliance on self-reported answers and cross-sectional nature.^4,7-10^ Given the proliferation of administrative claims and electronic health record (EHR) databases, there are now alternative sources to assess cancer screening utilization over time using historical billing and medical record information instead of self-reported information.^4,11^ The present study will examine administrative claims data to observe trends in breast, cervical, and colorectal cancer screening utilization from 2010 to 2019 by age group, geographic region, and test modality.

## Methods

### Study Sample

This retrospective, serial cross-sectional analysis utilized data from the Merative Health MarketScan Commercial Database from January 1, 2010, through December 31, 2019, encompassing 15.2 to 37.5 million enrollees with 12-month continuous enrollment in each calendar year during this period. The MarketScan commercial database is a nationwide convenience sample containing individual-level, de-identified healthcare claims information from employer-sponsored private health insurance plans. The commercial database contains the pooled healthcare experience of 141.2 million unique enrollees from 2010 to 2019 from multiple geographically dispersed states. The reporting of this study conforms to the STROBE guidelines for observational research.^12^ All enrollees meeting eligibility criteria were included annually; no sampling was performed. Data are fully de-identified; the study did not involve identifiable human subjects and was determined exempt by the Johns Hopkins University Institutional Review Board (IRB00020100).

### Measures and Outcomes

Cancer screening tests were identified using Current Procedural Terminology (CPT) and Healthcare Common Procedural Coding System (HCPCS) codes during the period from January 1, 2010, to December 31, 2019. The criteria for identifying excluded enrollees were defined using International Classification of Diseases (ICD-9/ICD-10) diagnosis and procedure codes. (Table S7, S9, S11) Annual screening rates were estimated to assess temporal trends in cancer screening over time rather than up-to-date screening (proportions of patients receiving screening within the recommended interval time). Up-to-date screening requires continuous enrollment across multi-year intervals.

Breast cancer screening rates were computed as the annual proportion of women who received screening mammography among eligible women aged 40 to 64 and continuously enrolled in a health insurance plan for 12 months each year.^13^ Rates are reported by age subgroup. We highlight women aged 50 to 64 as the group for whom screening has been most consistently recommended and include women aged 40 to 49 because recommendations for this range have differed across organizations and over time.^14,15^ Breast cancer screening test was identified using procedure codes for screening mammography.^16-19^ (Table S8) Diagnostic mammography was also captured among eligible women, as some physicians may bill for screening mammograms using diagnostic mammography codes.^16,20,21^ Eligible women with a prior history of breast cancer, breast involvement, breast signs or symptoms, and prior mastectomy were excluded.^18,19,22^ (Table S9)

Cervical cancer screening rates were computed as the annual proportion of women who receive (1) cytology test alone among eligible women aged 21 to 64 in a given year, (2) HPV test alone among eligible women aged 30 to 64 in a given year, or (3) co-testing among eligible women aged 30 to 64 in a given year.^5^ Co-testing was defined as cytology plus HPV testing within 3 days before or 30 days after the date of cytology test in a given year.^4^ Cervical cancer screening test was identified using procedure codes for cervical cytology, HPV testing, or co-testing.^4,17,23,24^ (Table S6) Women with a history of precancer or invasive cervical cancer or total hysterectomy were excluded. ^4,23,24^ (Table S7)

Colorectal cancer screening rates were computed as the annual proportion of patients who receive any of the following test modalities: (1) Guaiac fecal occult blood test (gFOBT) alone; (2) fecal immunochemical test (FIT) alone; (3) multitarget stool DNA with FIT component (sDNA-FIT); (4) Colonoscopy alone; (5) CT colonography alone; (6) flexible sigmoidoscopy (FS) alone; or (7) flexible sigmoidoscopy with FIT (FS+FIT) among the eligible population in a given year.^25^ We additionally report utilization among enrollees below screening age (0-44 years) and an overall 0-64 summary, alongside the primary 45-64 screening-eligible rate. Colorectal cancer screening tests were identified using relevant procedure codes.^17,22,26,27^ (Table S10) Eligible enrollees with a history of colorectal cancer and total colectomy were excluded.^22,27^ (Table S11)

Results were stratified by age, gender, region [metropolitan (urban) vs. non-metropolitan areas (rural), five US census regions], and type of health plan. The analysis was limited to 2010-2019 due to substantial disruptions in cancer screening during the COVID-19 pandemic in 2020. ^28-30^ The median and mean out-of-pocket (OOP) costs, including copay, coinsurance, and deductible, were estimated for each test modality across all years. Changes in screening utilization rates were computed as the percentage difference between 2010 and 2019 divided by the 2010 percentage.

### Statistical analysis

Annual screening rates were calculated as the number of eligible enrollees receiving a given test in a calendar year divided by the number of eligible enrollees in that year. Temporal trends from 2010 to 2019 were assessed using two-sided Poisson regression, with the individual screening indicator modeled at the person-year level and calendar year entered as a continuous predictor. Individuals could contribute observations across multiple years; within-person correlation across years was not adjusted for in these models. Urban-rural disparities in screening receipt (binary outcome) for 2019 were examined using GEE with a Poisson distribution, log link, independence correlation structure, and robust standard errors; each individual contributed a single observation, so no offset or repeated-measures adjustment applied. Covariates included sex (for the colorectal cohort), age group, place of residence (urban, rural, or unknown), U.S. census region, and health plan type. Enrollees with unknown place of residence or region were retained as a separate category rather than excluded. No a priori sample size calculation was performed, as the analysis included all eligible enrollees rather than a sample. Statistical significance was set at a two-sided alpha of 0.05. Analyses were performed using SAS 9.4 (SAS Institute).

## Results

### Description of the Sample

The study population was composed of enrollees with 12 months of continuous enrollment in a given claim year (Figure 1), ranging from 15.2 million in 2019 to 37.5 million in 2010, totaling 81.6 million unique enrollees from 2010 through 2019, after exclusion criteria are applied. (Table S4) Among the study population, 28.2% were younger than age 21 years, 28.7% were aged 21-39 years, and 43.2% were aged 40-64 years old. (Table 1) Enrollees aged 65 years or older were not included due to a lack of Medicare claims data availability. Approximately 82.2% of enrollees resided in urban areas, 13.4% in rural areas, and 4.5% in unknown areas. The distribution of urban residence remained relatively stable from 2010 through 2019, with the reported proportion of enrollees residing in urban regions ranging from 77.5% to 85.7%. Approximately 18.9% resided in the Northeast, 22% in North Central, 37.9% in the South, and 19.1% in the West. The regional distribution of enrollees was similar to the national population distribution reported by the U.S. Census Bureau.^31^

**Figure 1.**
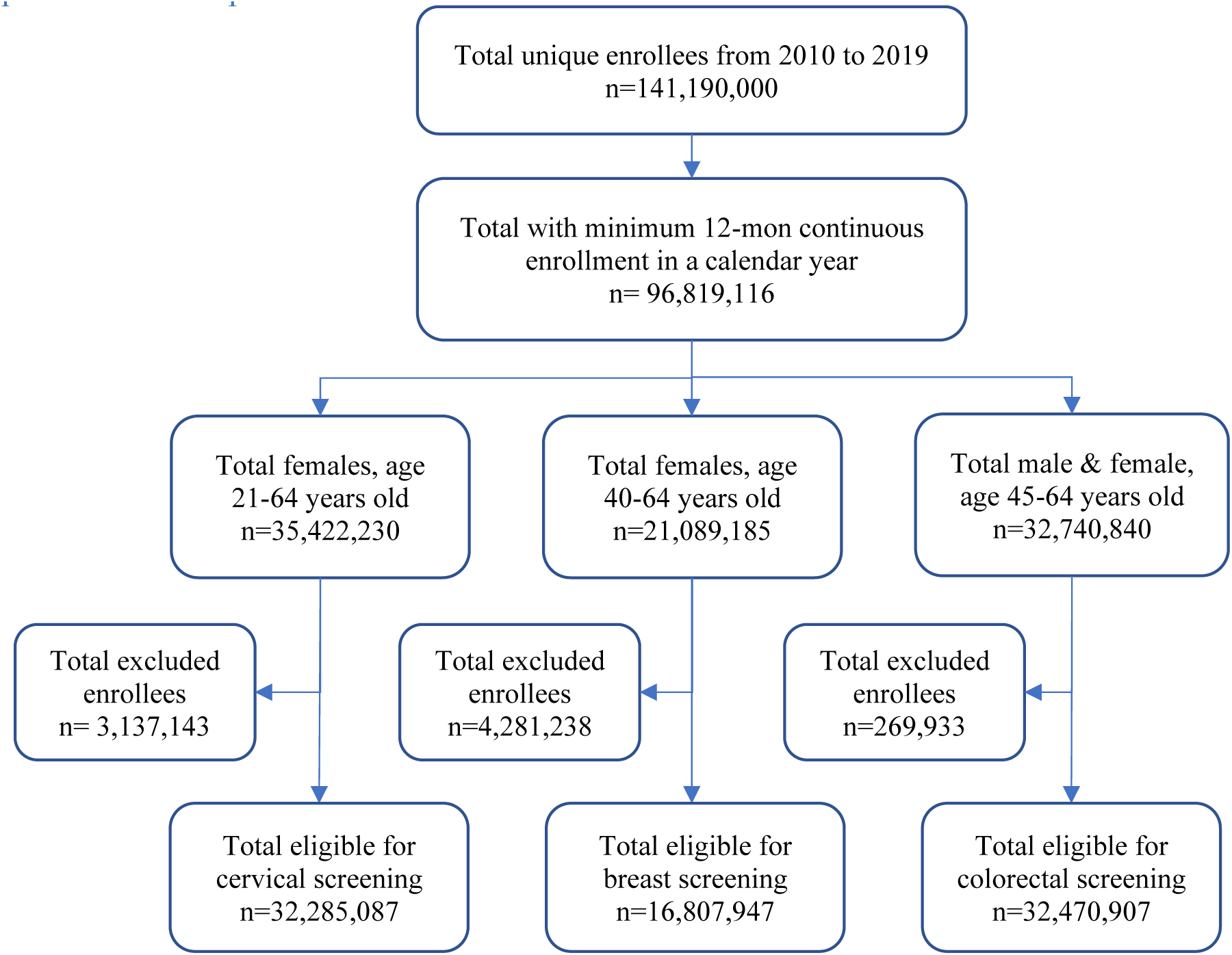
Study Population Flow Diagram.

**Table 1.** Baseline Demographics and Characteristics by Recommended Screening Cohorts.

| All Years (2010-2019) |  |  |  |  |
| --- | --- | --- | --- | --- |
| Variables | Cervical screening<br>Female,<br>age 21-64<br>years | Breast screening<br>Female,<br>age 40-64<br>years | Colorectal screening<br>Male/Female,<br>age 45-64<br>years | Overall Population<br>Male/Female, age<br>0-64 years |
| Total unique enrollees, n (%) | 32,285,087 | 16,807,947 | 32,470,907 | 81,563,941 (100%) |
| Female, % | 100% | 100% | 52.8% | 51.4% |
| Age Group, % |  |  |  |  |
| <21 years | - | - | - | 28.2% |
| 21-24 years | 8.2% | - | - | 7.3% |
| 25-29 years | 8% | - | - | 6.6% |
| 30-39 years | 20.9% | - | - | 14.8% |
| 40-44 years | 11.9% | 18.9% | - | 8.1% |
| 45-49 years | 13.1% | 20.8% | 24.4% | 8.6% |
| 50-54 years | 14% | 22.2% | 25.6% | 9% |
| 55-59 years | 13.7% | 21.8% | 24.9% | 8.7% |
| 60-64 years | 10.2% | 16.3% | 25.2% | 8.8% |
| Place of Residence, % |  |  |  |  |
| Unknown | 3.9% | 3.9% | 4.5% | 4.5% |
| Rural | 13.1% | 13.9% | 14.6% | 13.4% |
| Urban | 83% | 82.1% | 80.8% | 82.2% |
| Regions, % |  |  |  |  |
| Northeast | 18.2% | 18.8% | 20.1% | 18.9% |
| North Central | 21.3% | 21.5% | 21.8% | 22% |
| South | 40.8% | 40.6% | 38.1% | 37.9% |
| West | 18.3% | 17.8% | 18% | 19.1% |
| Unknown | 1.4% | 1.4% | 2% | 2.1% |
| Health Plan Type, % |  |  |  |  |
| Comprehensive/Basic major medical | 2.0% | 2.6% | 2.2% | 1.4% |
| PPO/EPO | 59.4% | 59.1% | 60.5% | 60.7% |
| HMO | 12.1% | 12% | 10.4% | 10.7% |
| CDHP/HDHP | 14.1% | 13.5% | 12.4% | 13.6% |
| POS/PSC | 7.6% | 7.6% | 7.9% | 7.6% |
| Unknown | 4.9% | 5.1% | 6.6% | 6.1% |

PPO: Preferred provider organization; EPO: Exclusive Provider Organization; HMO: Health Maintenance Organization; HDHP: High-deductible health plan; CDHP: Consumer-directed health plan; POS: Point of service, non-capitated; PSC: Point of service, capitated or partially capitated. Note: Individuals may appear in more than one cohort.

### Cervical Cancer Screening Utilization

Among women aged 21-29, cytology alone decreased from 43.1% in 2010 to 26.2% in 2019, a 39.2% decrease (p<0.001, Figure S2; Table 2). Among women aged 30-64, cytology alone declined 68.8%, from 28.2% in 2010 to 8.8% in 2019 (p<0.001, Figure S3; Table 2) while co-testing increased 78.1% from 11.4% in 2010 to 20.3% in 2019 (p<0.001). HPV testing alone remained low at approximately 0.2% annually from 2010 to 2019. Cervical cancer screening among women aged <21 years appropriately decreased from 2010 to 2019 (p<0.001, Figure S1), with only 0.7% receiving any cervical screening in 2019. (Table 2) Overall, any cervical cancer screening rates declined across all age groups between 2010 and 2019 (Figure S4).

**Table 2.**
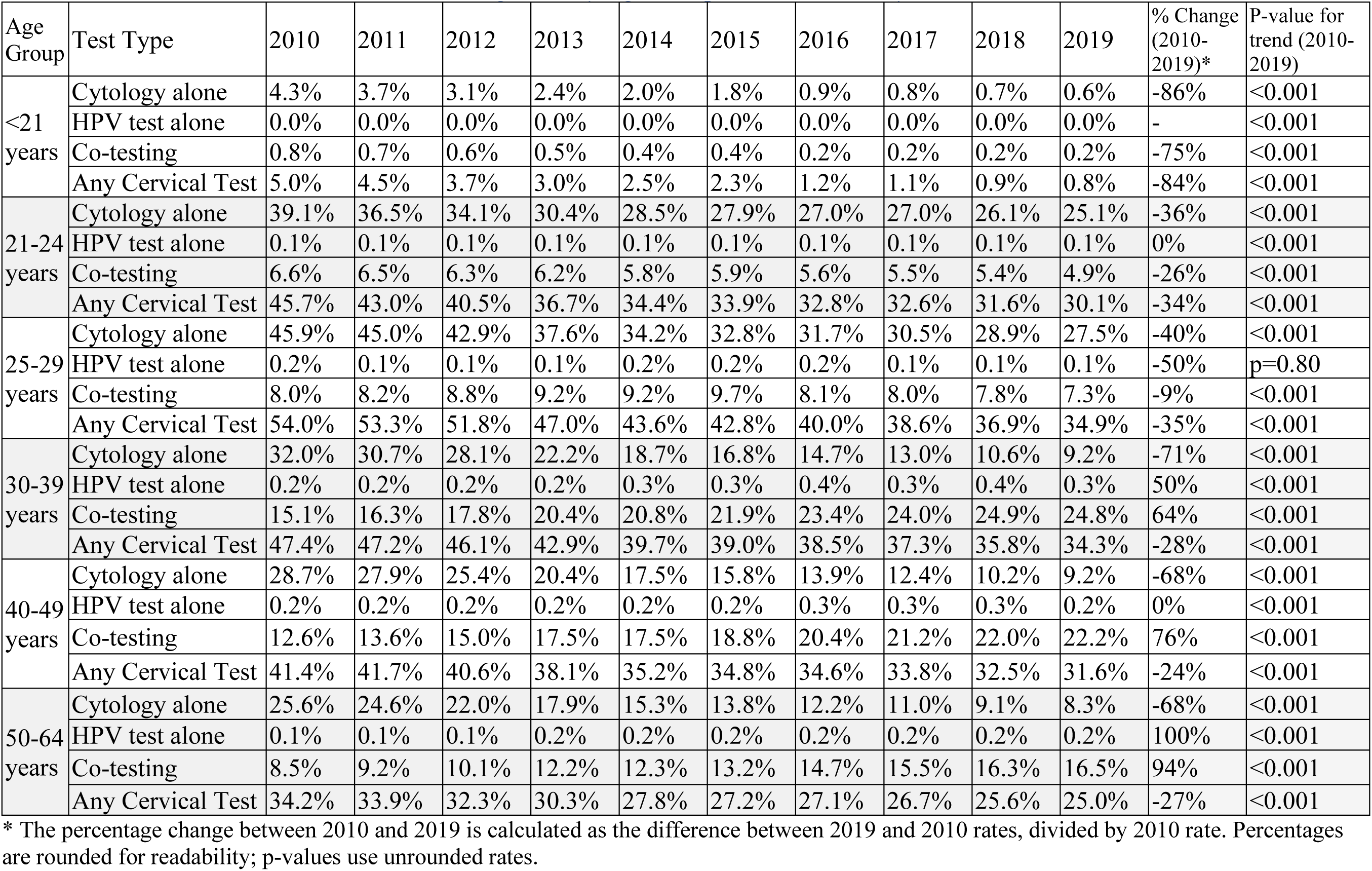
Annual Cervical Cancer Screening Rates by Age Group and Test Modality, 2010-2019.

In 2019, urban-residing women aged 21-64 had a significantly higher screening rate of cervical cancer screening compared with rural-residing counterparts (30.63% urban vs. 26.47% rural, absolute difference 4.17% [95% confidence interval [CI] 4.04-4.29]; IRR=1.04, 95% CI=1.04-1.05, p<0.001; Table 5, S5). Screening rates were highest in the Northeast and South census regions and lowest in the West (Table 5). Cervical cancer screening rates were highest in Alabama, New Jersey, Louisiana, New York, and Florida (25.7%-28.2%), and lowest in Utah, Wyoming, North Dakota, New Mexico, and South Dakota (12.6%-14.3%) (Figure 3). The median out-of-pocket costs for cytology, HPV testing alone, and co-testing was $0 throughout all years (Table S1).

### Breast Cancer Screening Utilization

Among women aged 40-49, screening mammography remained relatively stable, decreasing from 38.0% in 2010 to 37.4% in 2019 (Figure S5; Table 3). Among women aged 50-64 years, screening mammography rates remained stable at 45.7% in 2010 and 45.8% in 2019 (Figure S5; Table 3). Diagnostic mammography remained stable at approximately 5.8% for women aged 40-49 and 5.1% for women aged 50-64 from 2010 to 2019 (Figure S6; Table 3). Overall, the proportion of women aged <64 receiving any screening mammography declined from 19.6% in 2010 to 16.1% in 2019 (Figure S7). Screening mammography among women aged <40 appropriately decreased by 69% from 2010 to 2019 (p<0.001; Table 3).

**Table 3.** Annual Breast Cancer Screening and Diagnostic Mammography Rates by Age Group and Test Type, 2010-2019.

| Age Group | Test Type | 2010 | 2011 | 2012 | 2013 | 2014 | 2015 | 2016 | 2017 | 2018 | 2019 | % Change (2010-2019)* | P-value for trend (2010-2019) |
| --- | --- | --- | --- | --- | --- | --- | --- | --- | --- | --- | --- | --- | --- |
| <40 years | Screening Mammography | 1.6% | 1.4% | 1.3% | 1.2% | 1.1% | 1.1% | 0.6% | 0.6% | 0.6% | 0.5% | -69% | <0.001 |
|  | Diagnostic Mammography | 0.3% | 0.3% | 0.3% | 0.2% | 0.2% | 0.3% | 0.2% | 0.2% | 0.1% | 0.1% | -67% | <0.001 |
| 40-44 years | Screening Mammography | 35.6% | 36.7% | 36.6% | 36.8% | 36.1% | 36.1% | 33.6% | 34.0% | 34.2% | 34.9% | -2% | <0.001 |
|  | Diagnostic Mammography | 4.7% | 5.4% | 5.8% | 5.9% | 6.1% | 6.2% | 6.0% | 5.9% | 5.6% | 5.7% | 21% | <0.001 |
| 45-49 years | Screening Mammography | 40.1% | 41.0% | 40.7% | 41.2% | 40.1% | 40.5% | 39.0% | 39.3% | 39.4% | 39.8% | -1% | <0.001 |
|  | Diagnostic Mammography | 4.9% | 5.4% | 5.7% | 5.9% | 6.1% | 6.2% | 6.2% | 6.1% | 5.7% | 5.7% | 16% | <0.001 |
| 50-54 years | Screening Mammography | 43.4% | 43.8% | 43.2% | 43.9% | 42.7% | 43.5% | 42.6% | 43.5% | 43.8% | 44.2% | 2% | p=0.80 |
|  | Diagnostic Mammography | 4.6% | 5.0% | 5.2% | 5.3% | 5.5% | 5.7% | 5.7% | 5.7% | 5.5% | 5.5% | 20% | <0.001 |
| 55-59 years | Screening Mammography | 46.0% | 46.0% | 45.1% | 45.6% | 44.5% | 44.8% | 44.1% | 44.7% | 45.1% | 45.7% | -1% | <0.001 |
|  | Diagnostic Mammography | 4.4% | 4.6% | 4.7% | 4.9% | 5.0% | 5.1% | 5.2% | 5.2% | 4.9% | 4.9% | 11% | <0.001 |
| 60-64 years | Screening Mammography | 49.0% | 48.8% | 47.7% | 48.1% | 47.1% | 46.9% | 46.0% | 46.4% | 46.6% | 47.7% | -3% | <0.001 |
|  | Diagnostic Mammography | 4.6% | 4.8% | 4.9% | 5.1% | 5.1% | 5.2% | 5.2% | 5.2% | 4.9% | 4.9% | 7% | <0.001 |

\* The percentage change between 2010 and 2019 is calculated as the difference between 2019 and 2010 rates, divided by 2010 rate. Percentages are rounded for readability; p-values use unrounded rates.

In 2019, urban-residing women aged 40-64 had a significantly higher screening rate of screening mammography compared with rural-residing counterparts (42.67% urban vs. 41.48% rural, absolute difference 1.20% [95% CI 1.00-1.39], IRR=1.02, 95% CI=1.01-1.03, p<0.001; Table 5, S5). Screening mammography rates were highest in the North Central census region and lowest in the West (Table 5). Screening mammography rates were highest in Hawaii, Rhode Island, New Hampshire, Maine, and Louisiana (19.0%-21.4%), and lowest in Wyoming, Utah, Nevada, Alaska, and Idaho (9.4%-12.1%) (Figure 2). The median out-of-pocket cost for screening mammography was $0 across all years (Table S2).

**Figure 2.**
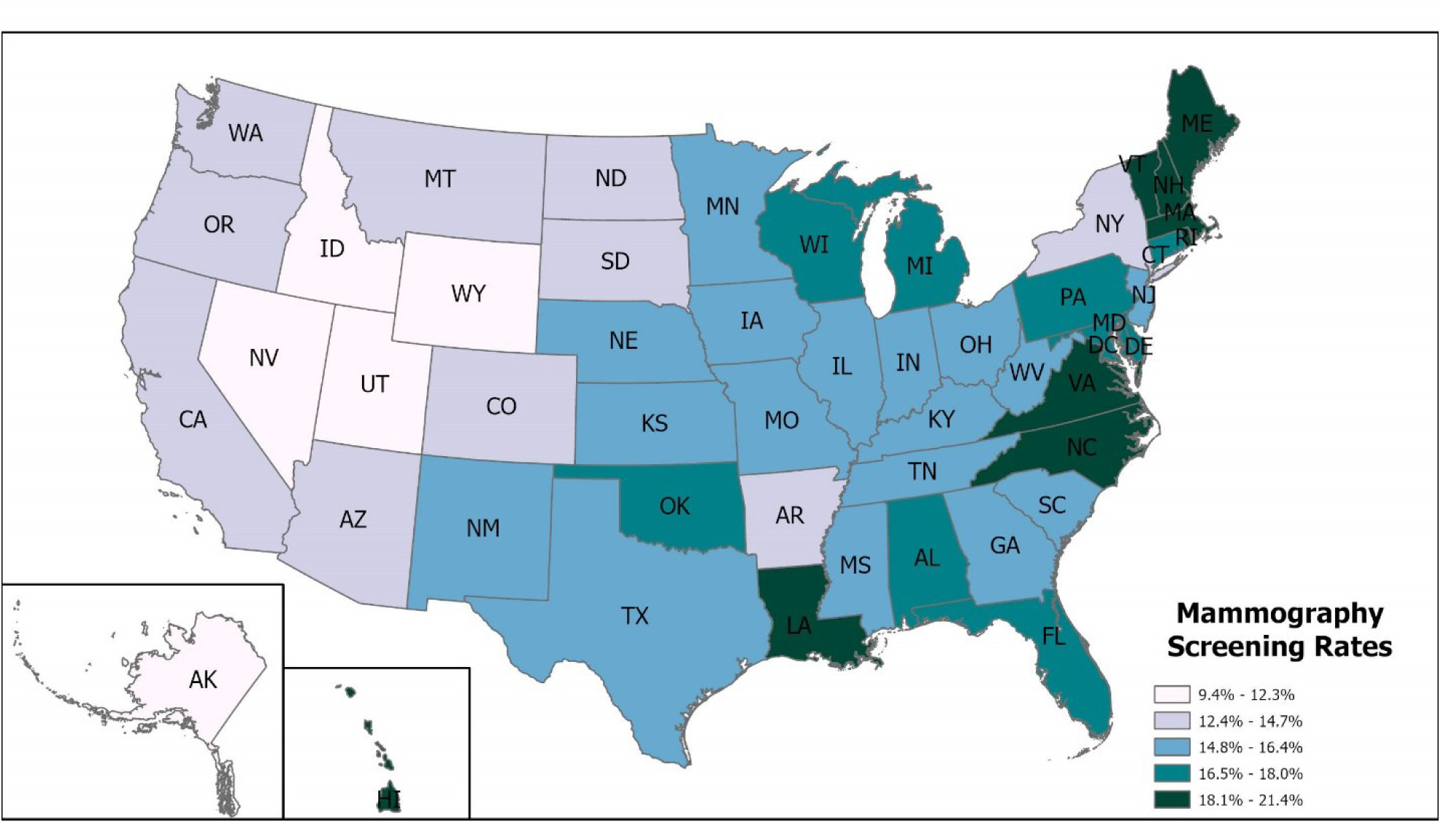
Screening Mammography Rates by U.S. States in 2019.

**Figure 3.**
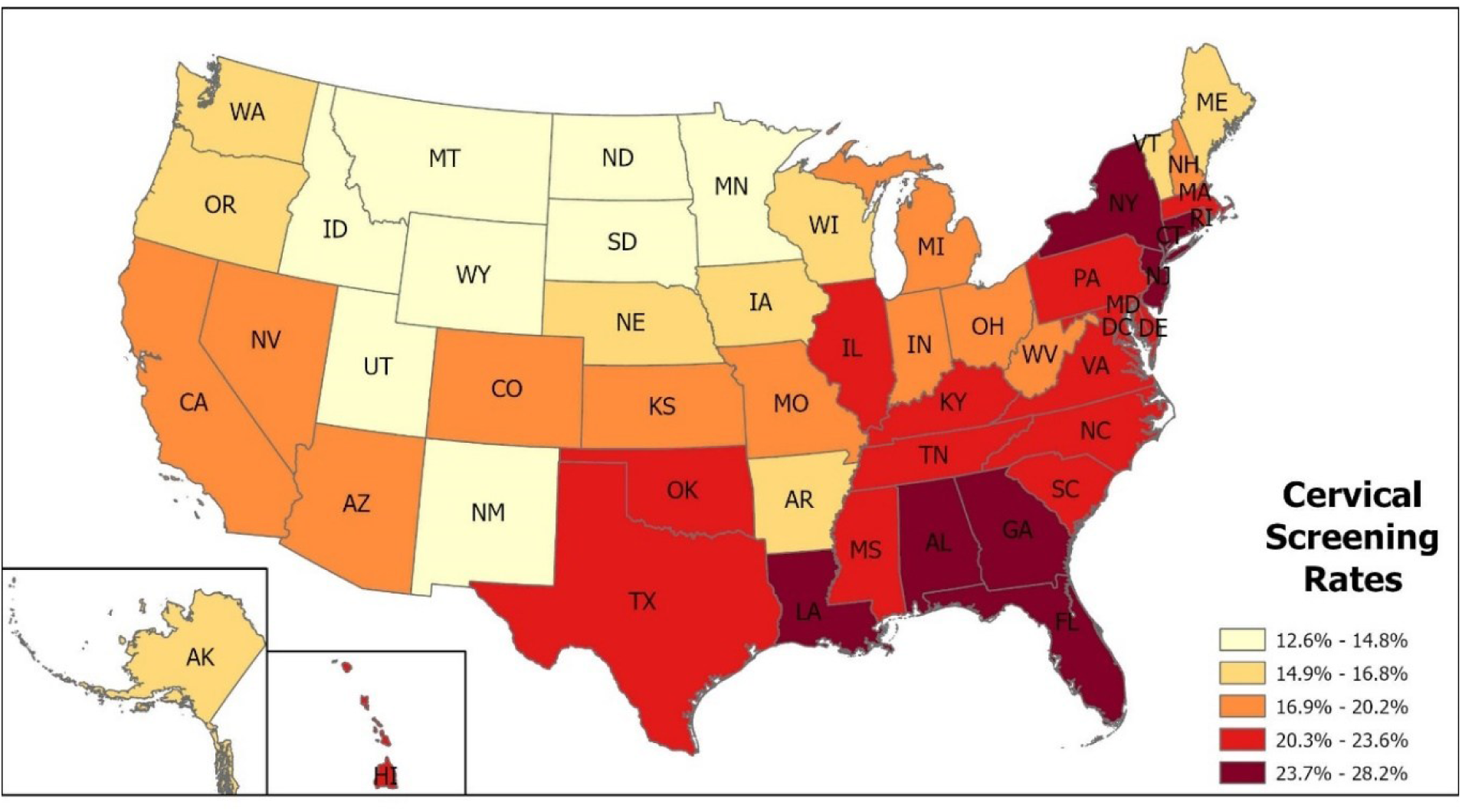
Cervical Screening Rates by U.S. States in 2019.

### Colorectal Cancer Screening Utilization

Among adults aged 45-49 years, any colorectal cancer screening decreased from 11.2% in 2010 to 7.0% in 2019 (Figures S8, S9; Table 4). Among adults aged 50-64, screening remained relatively stable, decreasing from 19.6% in 2010 to 18.6% in 2019 (Figures S8, S9; Table 4). Screening among enrollees aged 0-44 appropriately decreased by 36% from 2010 to 2019 (p<0.001; Figures S8, S9; Table 4). Among adults aged 45-49, utilization of specific screening modalities declined significantly between 2010 and 2019, with flexible sigmoidoscopy declining by 50%, colonoscopy by 15%, FOBT by 74%, FIT by 27%, and DCBE by 100% (Figure S10; Table 4). Similar but less severe declines were observed among adults aged 50-64 (Figure S11; Table 4). Among all age groups (0-64 years), colonoscopy utilization slightly increased from 3.6% to 3.8%, and FIT utilization remained stable between 1.4% and 1.7%. Flexible sigmoidoscopy and FOBT declined substantially. FIT-DNA utilization increased from 0.0% in 2015 to 0.4% in 2019 (Figure S12; Table 4). Overall, any colorectal cancer screening across enrollees aged 0 to 64 (including those below screening age) decreased from 7.7% in 2010 to 6.5% in 2019 (Figure S12).

**Table 4.** Annual Colorectal Cancer Screening Test by Age Group and Test Modality, 2010-2019.

| Age Group | Test Type | 2010 | 2011 | 2012 | 2013 | 2014 | 2015 | 2016 | 2017 | 2018 | 2019 | % Change (2010-2019)* | P-value for trend (2010-2019) |
| --- | --- | --- | --- | --- | --- | --- | --- | --- | --- | --- | --- | --- | --- |
| 0-44 years | Flexible Sigmoidoscopy | 0.1% | 0.1% | 0.1% | 0.1% | 0.1% | 0.1% | 0.1% | 0.1% | 0.1% | 0.1% | 0% | <0.001 |
|  | Colonoscopy | 0.9% | 0.9% | 0.9% | 0.8% | 0.8% | 0.8% | 0.8% | 0.8% | 0.8% | 0.8% | -11% | <0.001 |
|  | CT Colonography | 0% | 0% | 0% | 0% | 0% | 0% | 0% | 0% | 0% | 0% | - | <0.05 |
|  | FOBT | 0.8% | 0.7% | 0.6% | 0.5% | 0.5% | 0.4% | 0.3% | 0.3% | 0.2% | 0.2% | -75% | <0.001 |
|  | FIT | 0.4% | 0.4% | 0.4% | 0.4% | 0.4% | 0.4% | 0.3% | 0.3% | 0.3% | 0.3% | -25% | <0.001 |
|  | DCBE | 0% | 0% | 0% | 0% | 0% | 0% | 0% | 0% | 0% | 0% | - | <0.001 |
|  | FIT-DNA | 0% | 0% | 0% | 0% | 0% | 0% | 0% | 0% | 0% | 0% | - | <0.001 |
|  | Any Colorectal Test | 2.2% | 2.1% | 2% | 1.9% | 1.8% | 1.7% | 1.5% | 1.5% | 1.5% | 1.4% | -36% | <0.001 |
| 45-49 years | Flexible Sigmoidoscopy | 0.2% | 0.2% | 0.2% | 0.2% | 0.2% | 0.2% | 0.2% | 0.2% | 0.1% | 0.1% | -50% | <0.001 |
|  | Colonoscopy | 4.8% | 4.8% | 4.7% | 4.8% | 4.8% | 4.8% | 3.6% | 3.6% | 3.8% | 4.1% | -15% | <0.001 |
|  | CT Colonography | 0% | 0% | 0% | 0% | 0% | 0% | 0% | 0% | 0% | 0% | - | <0.05 |
|  | FOBT | 3.9% | 3.5% | 3.1% | 2.8% | 2.5% | 2.1% | 1.7% | 1.5% | 1.2% | 1% | -74% | <0.001 |
|  | FIT | 2.2% | 2.6% | 2.6% | 2.6% | 2.3% | 2.3% | 2% | 1.9% | 1.7% | 1.6% | -27% | <0.001 |
|  | DCBE | 0.1% | 0.1% | 0% | 0% | 0% | 0% | 0% | 0% | 0% | 0% | -100% | <0.001 |
|  | FIT-DNA | 0% | 0% | 0% | 0% | 0% | 0% | 0% | 0% | 0% | 0.1% | - | <0.001 |
|  | Any Colorectal Test | 11% | 11% | 11% | 10% | 9.8% | 9.4% | 7.5% | 7.1% | 7% | 7% | -38% | <0.001 |
| 50-64 years | Flexible Sigmoidoscopy | 0.3% | 0.3% | 0.3% | 0.2% | 0.2% | 0.2% | 0.2% | 0.2% | 0.2% | 0.2% | -33% | <0.001 |
|  | Colonoscopy | 9.5% | 9.5% | 9.6% | 9.8% | 9.8% | 10% | 11% | 11% | 11% | 11% | 13% | <0.001 |
|  | CT Colonography | 0% | 0% | 0% | 0% | 0% | 0% | 0% | 0% | 0% | 0% | - | <0.001 |
|  | FOBT | 5.9% | 5.3% | 4.8% | 4.3% | 4.1% | 3.5% | 3.1% | 2.9% | 2.5% | 2.1% | -64% | <0.001 |
|  | FIT | 3.7% | 4.4% | 4.8% | 5.2% | 4.4% | 4.6% | 4.7% | 4.7% | 4.4% | 4.1% | 11% | <0.001 |
|  | DCBE | 0.1% | 0.1% | 0.1% | 0.1% | 0.1% | 0.1% | 0.1% | 0.1% | 0.1% | 0% | -100% | <0.001 |
|  | FIT-DNA | 0% | 0% | 0% | 0% | 0% | 0% | 0.1% | 0.3% | 0.6% | 1.5% | - | <0.001 |
|  | Any Colorectal Test | 20% | 20% | 20% | 20% | 19% | 19% | 19% | 19% | 19% | 19% | -5% | <0.001 |

\* The percentage change between 2010 and 2019 is calculated as the difference between 2019 and 2010 rates, divided by 2010 rate. Percentages are rounded for readability; p-values use unrounded rates. FOBT: Fecal Occult Blood Test; FIT: Fecal Immunochemical Test; DCBE: Double-Contrast Barium Enema; FIT-DNA: Fecal Immunochemical Test-Deoxyribonucleic Acid (DNA) (i.e., Cologuard); CT: Computerized Tomography

In 2019, urban-residing adults aged 45-64 had a significantly higher screening rate of any colorectal cancer screening compared with rural-residing counterparts (16.10% urban vs. 14.22% rural, absolute difference 1.88% [95% CI 1.79-1.97] IRR=1.02, 95% CI=1.01-1.03, p<0.001; Table 5, S5). Colorectal cancer screening rates were highest in the South census region and lowest in the North Central census region (Table 5). Colorectal cancer screening rates were highest in Hawaii, Rhode Island, Connecticut, Alabama, and Michigan (8.1%-10.5%), and lowest in Wyoming, Utah, South Dakota, Nevada, and Alaska (3.7%-3.9%) (Figure 4). The median out-of-pocket cost for any colorectal cancer screening test was $0 for all years (Table S3).

**Figure 4.**
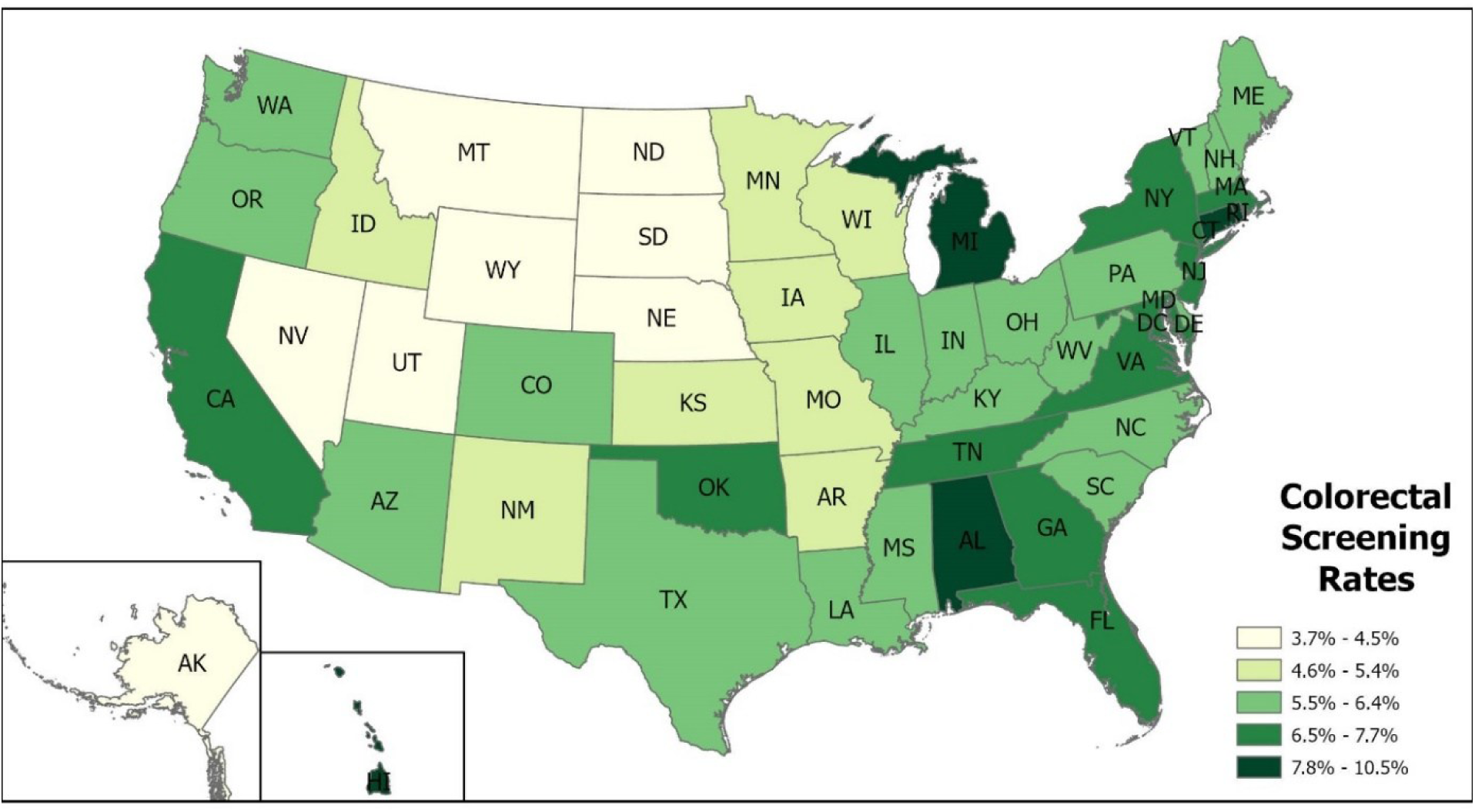
Colorectal Cancer Screening Rates by U.S. States in 2019.

**Table 5.** Multivariable Regression Results using Generalized Estimating Equations (GEE) with Poisson Distribution Models Predictors of Cancer Screening for 2019 Claim Year.

| Variables | Incidence Rate Ratio (IRR) |  |  |
| --- | --- | --- | --- |
|  | Cervical Cancer Screening | Breast Cancer Screening | Colorectal Cancer Screening |
| **Gender** |  |  |  |
| Female | - | - | Ref |
| Male | - | - | 0.86 (0.85 - 0.86)*** |
| **Age group** |  |  |  |
| 21-24 years | Ref | - | - |
| 25-29 years | 1.15 (1.14 - 1.16)*** | - | - |
| 30-39 years | 1.13 (1.13 - 1.14)*** | - | - |
| 40-44 years | 1.07 (1.06 - 1.08)*** | Ref | - |
| 45-49 years | 1.02 (1.01 - 1.03)*** | 1.14 (1.13 - 1.15)*** | Ref |
| 50-54 years | 0.93 (0.92 - 0.93)*** | 1.27 (1.26 - 1.28)*** | 2.75 (2.73 - 2.78)*** |
| 55-59 years | 0.82 (0.82 - 0.83)*** | 1.31 (1.30 - 1.32)*** | 2.40 (2.38 - 2.42)*** |
| 60-64 years | 0.73 (0.73 - 0.74)*** | 1.37 (1.36 - 1.38)*** | 2.84 (2.82 - 2.86)*** |
| **Place of Residence** |  |  |  |
| Rural | Ref | Ref | Ref |
| Urban | 1.05 (1.04 - 1.05)*** | 1.02 (1.01 - 1.03)*** | 1.02 (1.01 - 1.03)** |
| Unknown | 1.14 (1.13 - 1.15)*** | 1.05 (1.05 - 1.06)*** | 1.14 (1.14 - 1.15)*** |
| **US census region** |  |  |  |
| Northeast | Ref | Ref | Ref |
| North Central | 0.85 (0.85 - 0.86)*** | 1.06 (1.05 - 1.06)*** | 0.96 (0.95 - 0.97)*** |
| South | 1.00 (1.00 - 1.01) | 1.05 (1.04 - 1.06)*** | 1.06 (1.05 - 1.07)*** |
| West | 0.77 (0.77 - 0.78)*** | 0.89 (0.89 - 0.90)*** | 1.05 (1.05 - 1.06)*** |
| Unknown | 0.51 (0.49 - 0.53)*** | 0.36 (0.34 - 0.38)*** | 0.95 (0.91 - 0.99)* |
| **Type of health plan** |  |  |  |
| Comprehensive/Basic major medical | Ref | Ref | Ref |
| PPO/EPO | 1.10 (1.09 - 1.12)*** | 1.18 (1.16 - 1.20)*** | 0.98 (0.97 - 0.99)** |
| HMO | 1.07 (1.06 - 1.09)*** | 1.20 (1.18 - 1.22)*** | 1.12 (1.11 - 1.14)*** |
| POS/PSC | 1.16 (1.14 - 1.18)*** | 1.19 (1.17 - 1.21)*** | 1.00 (0.99 - 1.02) |
| CDHP/HDHP | 1.12 (1.10 - 1.14)*** | 1.21 (1.19 - 1.23)*** | 1.03 (1.01 - 1.04)*** |
| Unknown | 0.97 (0.95 - 0.99)*** | 1.21 (1.18 - 1.23)*** | 0.94 (0.92 - 0.96)*** |

PPO: Preferred provider organization; EPO: Exclusive Provider Organization; HMO: Health Maintenance Organization; HDHP: High-deductible health plan; CDHP: Consumer-directed health plan; POS: Point of service, non-capitated; PSC: Point of service, capitated or partially capitated. \* $p < 0.05$ ; \*\* $p < 0.01$ ; \*\*\* $p < 0.001$ .

## Discussion

### Cervical Cancer Screening

Cervical cancer screening patterns from 2010 to 2019 demonstrated appropriate alignment with evolving guidelines. Cytology alone declined substantially, particularly among women aged 30-64, while co-testing increased, reflecting guidelines from the USPSTF, ACOG, and ACS recommending co-testing every 5 years for women aged 30-65 since 2012.^4,32-34^ Screening among women aged <21 appropriately decreased, consistent with evidence that harms outweigh benefits in this age group.^5,35^ However, HPV testing alone remained underutilized at approximately 0.2% annually, despite its inclusion as an option by ACOG (2016), USPSTF (2018), and ACS (2020).^4,5,36,37^ Uptake may increase as more guidelines designate primary HPV testing as preferred.^4,38^

These findings align with other national studies. Analysis of 2005-2019 NHIS data showed the proportion of women without up-to-date screening increased from 14.4% to 23.0%, with lack of knowledge as the primary barrier.^39^ Similarly, another NHIS analysis reported that recent cervical cancer screening rates declined from 82.9% in 2008 to 80.0% in 2018.^40^ Geographic disparities observed in this study are consistent with published literature showing higher screening rates in urban versus rural areas and regional variations, with the Northeast reporting higher co-testing uptake.^39^ An NHIS analysis found rural residents had significantly higher rates of overdue screening compared with urban residents (26.2% vs. 22.6%, p<0.05).^39^ In this study, the absolute urban-rural difference was 4.17% in 2019, the largest gap among the three cancers examined. Given cervical cancer’s long preclinical window, this gap likely reflects a clinically meaningful disparity in urban-rural early detection opportunity.

### Breast Cancer Screening

Screening mammography rates remained stagnant or declined across all age groups from 2010 to 2019, despite well-established evidence supporting early detection benefits.^41^ This trend is consistent with other national studies showing declining mammography rates.^42-45^ Analysis of National Health Interview Survey (NHIS) data from 2000 to 2015 reported adjusted mammography rates declined by 3.0% (p<0.05).^42^ Particularly, rates among women aged 50-64, the highest-risk group, remained essentially unchanged at 45.7% in 2010 and 45.8% in 2019. Barriers to screening may include limited awareness or education,^46^ access to healthcare,^47,48^ fear and discomfort,^47,49,50^ and inconsistent physician recommendations.^48,51^

Significant geographic disparities were observed, with highest rates in the North Central region and lowest in the West. Urban-residing women aged 40-64 had higher screening rates compared with rural counterparts, consistent with published literature on urban-rural disparities.^52,53^ For example, analysis of 2012-2016 Behavioral Risk Factor Surveillance System (BRFSS) data found urban women had higher odds of screening mammography compared with rural women (Odds Ratio [OR]: 1.06, 95% CI 1.02-1.10, p<0.01).^52^ In this study, the absolute urban-rural difference was 1.20% in 2019, a comparatively modest gap relative to the other cancers examined, but clinically meaningful on a population-level scale. These findings point to potential targets for interventions to improve screening uptake, particularly in rural areas and underserved populations. As a broader implication, state-level policies such as Medicaid expansion under the Affordable Care Act (ACA) may help reduce geographic disparities and expand access to care in the wider population, although this possibility extends beyond the commercially insured population studied here.

### Colorectal Cancer Screening

Colorectal cancer screening utilization declined modestly among commercially insured enrollees from 2010 to 2019, with a more pronounced decline among adults aged 45-49. The decline among individuals under age 50 is consistent with 2008 USPSTF guidelines recommending screening initiation at age 50. However, the declining trend among adults aged 50-64, though less severe, is concerning given increasing cancer risk with age. Among adults aged 50-64, colonoscopy and fecal immunochemical test (FIT) remained relatively stable, suggesting established patient and physician preferences for these modalities. In contrast, flexible sigmoidoscopy, guaiac fecal occult blood test (FOBT), and double-contrast barium enema (DCBE) declined substantially. The emergence of multitarget stool DNA with FIT component (FIT-DNA) from 0.0% in 2015 to 0.4% in 2019 represents a new screening option, though uptake remains low.

Significant urban-rural disparities were observed, with urban-residing adults aged 45-64 having significantly higher screening rates than rural counterparts in 2019. These findings align with published literature.^54-56^ Analysis of 2014-2019 Behavioral Risk Factor Surveillance System (BRFSS) data found colorectal cancer screening rates were 4.4% higher in urban regions (72.8%) compared with rural regions (68.4%, p<0.001).^56^ In the present study, the absolute urban-rural difference was 1.88% in 2019, smaller than the BRFSS estimate, but clinically meaningful at the scale of this population. Improved access to preventive screening services in rural areas is needed to promote early detection and reduce cancer-related mortality.

### Strengths and Limitations

This study’s primary strength is the large sample size of approximately 141.2 million unique enrollees from geographically diverse regions mirroring the U.S. population distribution. However, several limitations exist. The MarketScan commercial claims database is a convenience sample of employer-sponsored insured enrollees and their dependents, excluding Medicare, Medicare Advantage, Medicaid beneficiaries, and uninsured individuals. The database does not include race/ethnicity or socioeconomic measures such as income, education, and occupation.

Exclusion of adults aged 65 and older due to lack of Medicare claims availability creates a data gap for the 65-75 age group, limiting generalizability for colorectal and breast cancer screening cohorts where guidelines recommend screening through age 75. This study assessed annual screening rates rather than adherence to recommended screening intervals (up-to-date screening). Administrative claims data are susceptible to miscoding, data entry errors, and missing data. Mammography coded as diagnostic may reflect symptomatic or follow-up imaging rather than screening, and colonoscopy may be performed for screening, surveillance, or diagnostic purposes. Such misclassification may lead to under- or over-counting of screening events. Finally, the large sample size may yield statistically significant results for clinically small changes in screening rates; findings should be interpreted considering both statistical significance and clinical relevance. For this reason, we report absolute differences in screening rates alongside incidence rate ratios and interpret small differences cautiously even when they reach statistical significance.

## Conclusion

Despite consensus on the benefits of early cancer detection, utilization of cervical, breast, and colorectal cancer screening among commercially insured adults remained stagnant or declined from 2010 to 2019 in the US. For cervical cancer, cytology alone declined while co-testing increased among women aged 30-64, appropriately reflecting guideline changes. However, HPV testing alone remained underutilized. Screening mammography rates were persistently low across all age groups, particularly concerning for women aged 50-64 at highest risk. Colorectal cancer screening declined modestly, with pronounced decreases among adults aged 45-49.

Persistent urban-rural disparities were observed across all three cancer types, with urban residents consistently demonstrating higher screening rates. These findings suggest that targeted interventions may help improve screening utilization, particularly in rural and underserved populations. Strategies to address barriers including access to care, patient and provider education, and policy interventions such as Medicaid expansion may help reduce geographic disparities and improve cancer screening uptake.

## Informed consent statement

Informed consent was not applicable to this study, as all data were derived from MarketScan commercial claims database, a fully de-identified, HIPAA-compliant secondary dataset.

## Ethics approval statement

The Johns Hopkins University Institutional Review Board (IRB) (IRB00020100) determined this study to be exempt from IRB oversight.

## Data availability statement

The data underlying the results presented in the study are available from the Merative MarketScan Commercial Claims and Encounters Database. Data cannot be shared publicly because of data use agreement restrictions and licensing limitations. MarketScan data are available for licensing from Merative (formerly IBM Watson Health) at https://www.merative.com

## Abbreviations

ACA: Affordable Care Act
ACOG: American College of Obstetricians and Gynecologists
ACS: American Cancer Society
BRFSS: Behavioral Risk Factor Surveillance System
CI: confidence interval
CPT: Current Procedural Terminology
DCBE: double-contrast barium enema
EHR: electronic health record
FIT: fecal immunochemical test
FIT-DNA: multitarget stool DNA test
gFOBT: guaiac fecal occult blood test
GEE: generalized estimating equations
HCPCS: Healthcare Common Procedure Coding System
HINTS: Health Information National Trends Survey
HPV: human papillomavirus
ICD: International Classification of Diseases
IRR: incidence rate ratio
NHIS: National Health Interview Survey
OOP: out-of-pocket
USPSTF: United States Preventive Services Task Force.

