## Supplemental Materials for "Trends in the Utilization of Breast, Cervical, and Colorectal Cancer Screening from 2010 to 2019 Among a Commercially Insured Population Using the MarketScan Commercial Claims Database"

**List of Supplemental Figures**

Figure S1 Cervical Cancer Screening Rates by Testing Type, Age <21 ... 5

Figure S2 Cervical Cancer Screening Rates by Testing Type, Age 21-29 ... 5

Figure S3 Cervical Cancer Screening Rates by Testing Type, Age 30-64 ... 6

Figure S4 Cervical Cancer Screening Rates by Test Type, All Age Groups ... 6

Figure S5 Screening Mammography Rates by Age Groups ... 7

Figure S6 Diagnostic Mammography Rates by Age Groups ... 7

Figure S7 Overall Screening and Diagnostic Mammography Rates, All Age Groups ... 8

Figure S8 Any Colorectal Cancer Screening Rate by Age Group (A) ... 8

Figure S9 Any Colorectal Cancer Screening Rate by Age Group (B) ... 9

Figure S10 Colorectal Cancer Screening by Test Modality Type, Age 45-49 ... 9

Figure S11 Colorectal Cancer Screening by Test Modality Type, Age 50-64 ... 10

Figure S12 Colorectal Cancer Screening Rates by Test Modalities, All Age (0-64 years) ... 10

#### List of Supplemental Tables

Table S1 Mean and Median Out-of-Pocket (OOP) Costs by Test Modality Type for Cervical Cancer Screening 11

Table S2 Mean and Median Out-of-Pocket (OOP) Costs by Test Modality Type for Breast Cancer Screening 11

Table S3 Mean and Median Out-of-Pocket (OOP) Costs by Test Modality Type for Colorectal Cancer Screening 12

Table S4 Total Unique Patient Population by Claim Year, 2010-2019 ... 13

Table S5 2019 Unadjusted Cancer Screening Rates by Place of Residence ... 13

Table S6 List of Procedure and Diagnosis Codes for Cervical Cancer Screening, including Cytology and HPV Testing ... 14

### Supplemental Results

#### Cervical Cancer Screening Figures

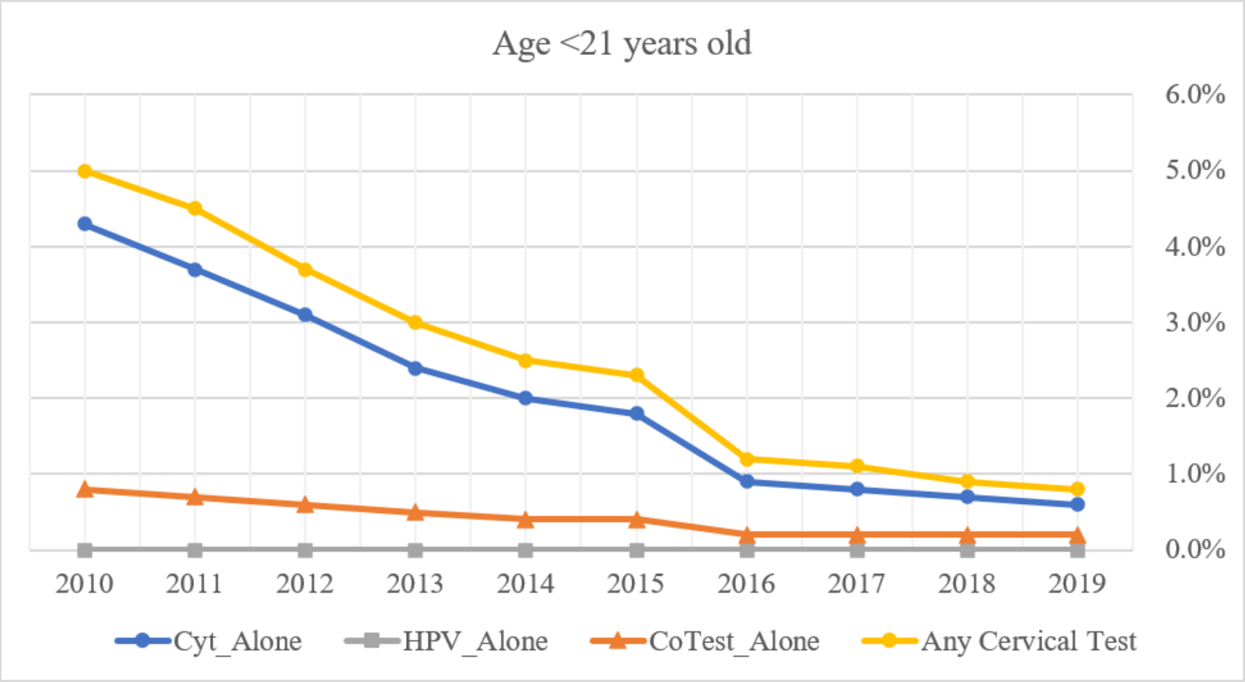

*Figure S1 Cervical Cancer Screening Rates by Testing Type, Age <21*

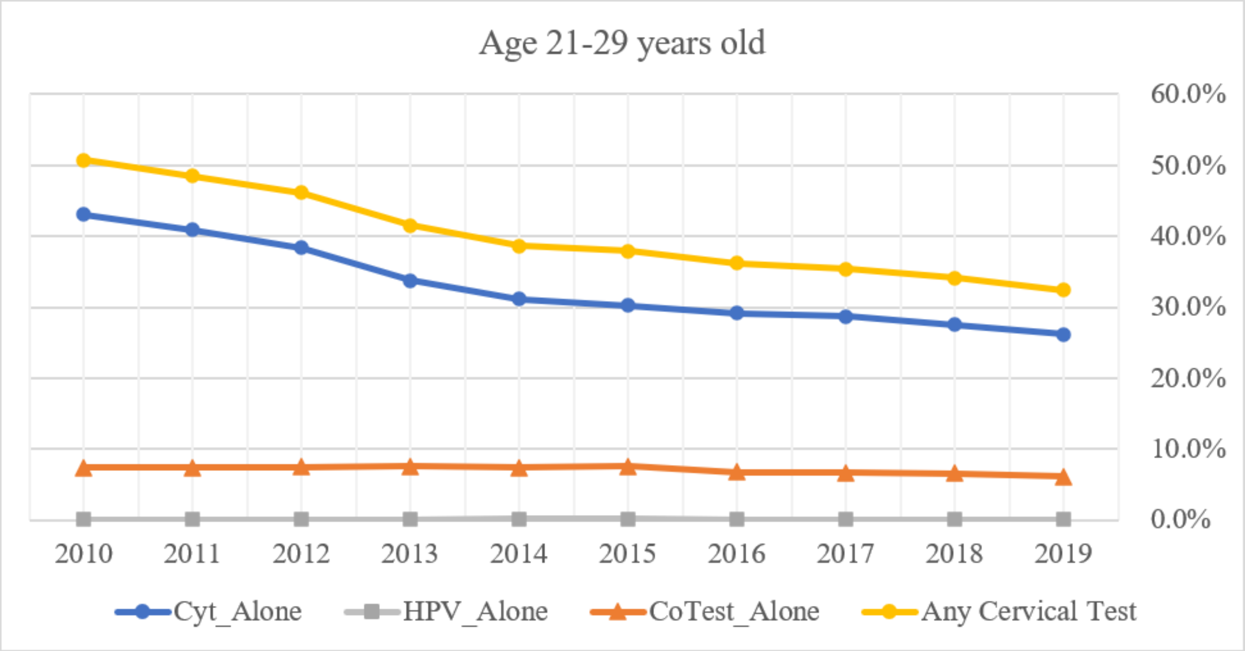

*Figure S2 Cervical Cancer Screening Rates by Testing Type, Age 21-29*

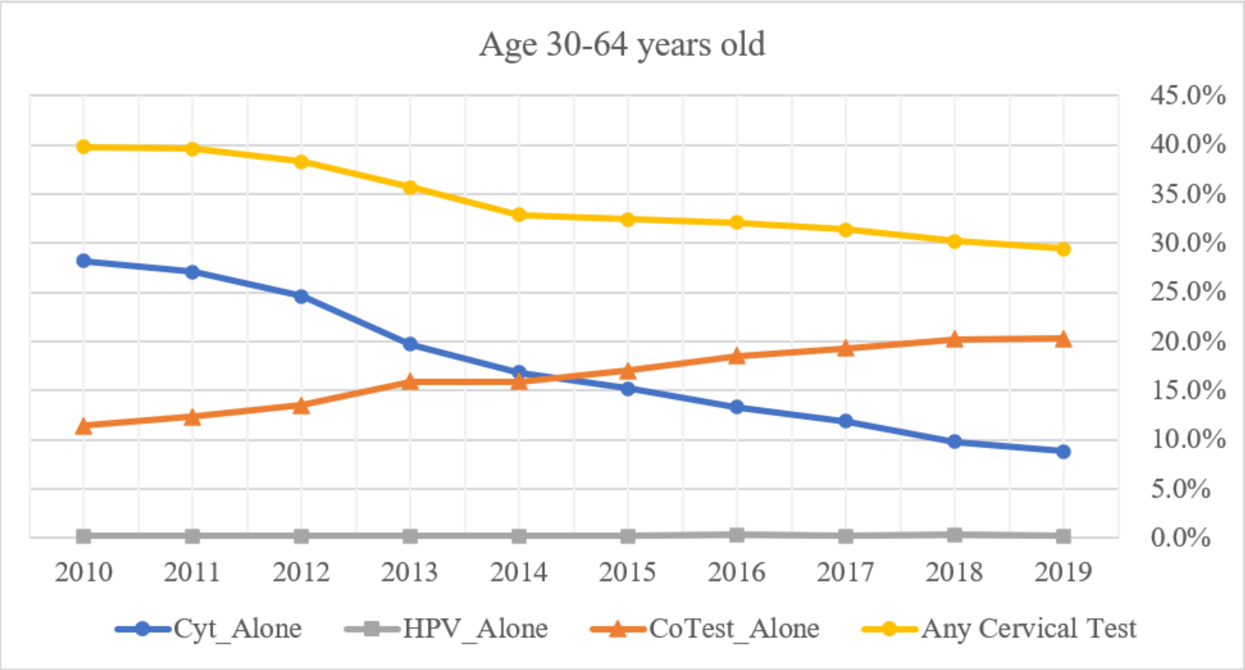

*Figure S3 Cervical Cancer Screening Rates by Testing Type, Age 30-64*

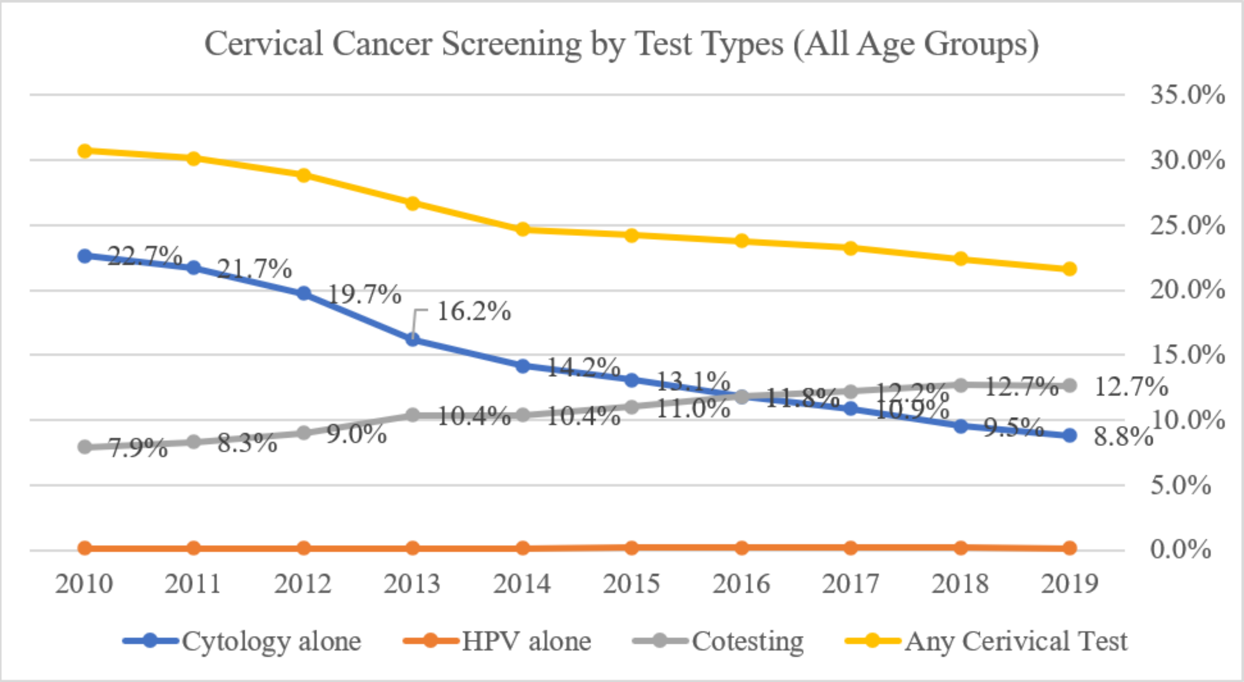

*Figure S4 Cervical Cancer Screening Rates by Test Type, All Age Groups*

#### Breast Cancer Screening Figures

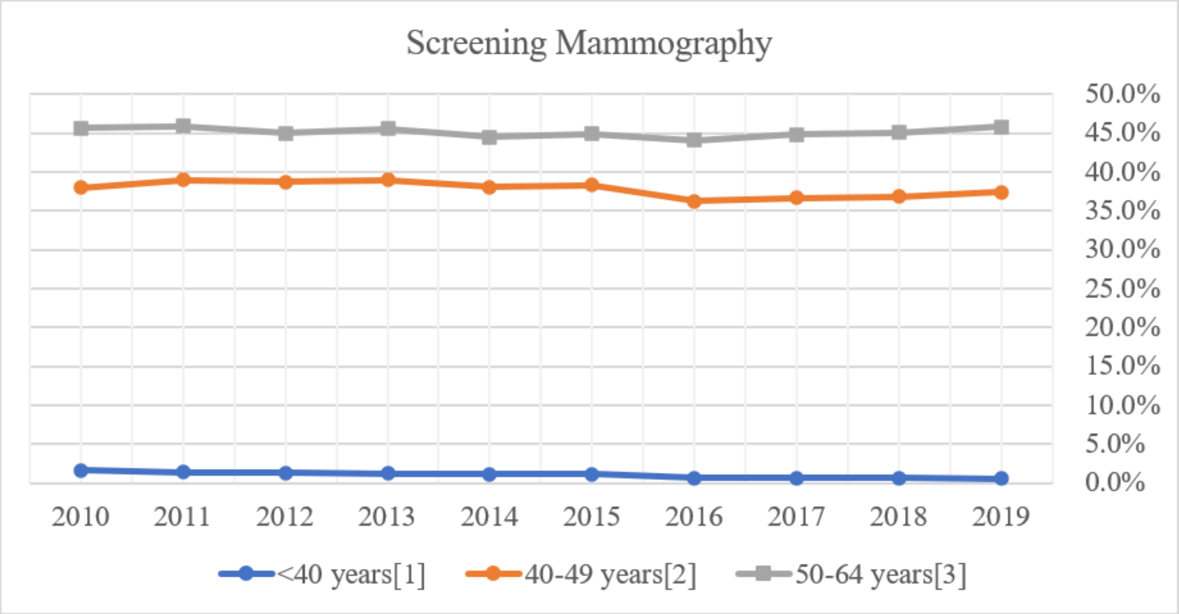

*Figure S5 Screening Mammography Rates by Age Groups*

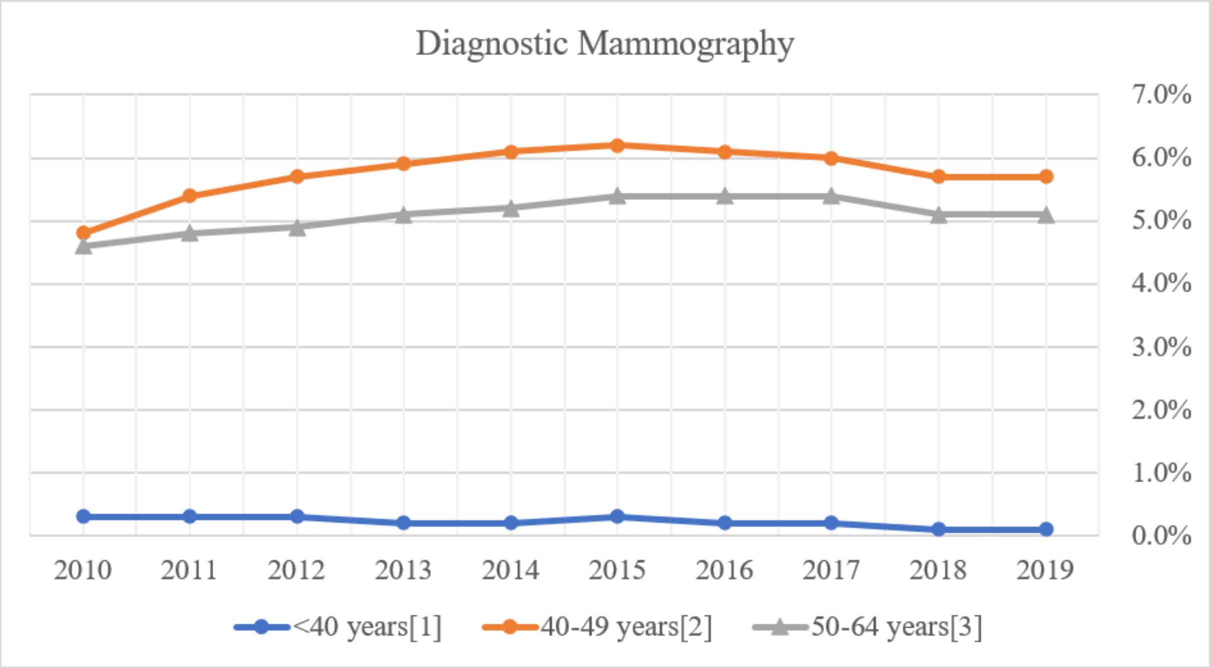

*Figure S6 Diagnostic Mammography Rates by Age Groups*

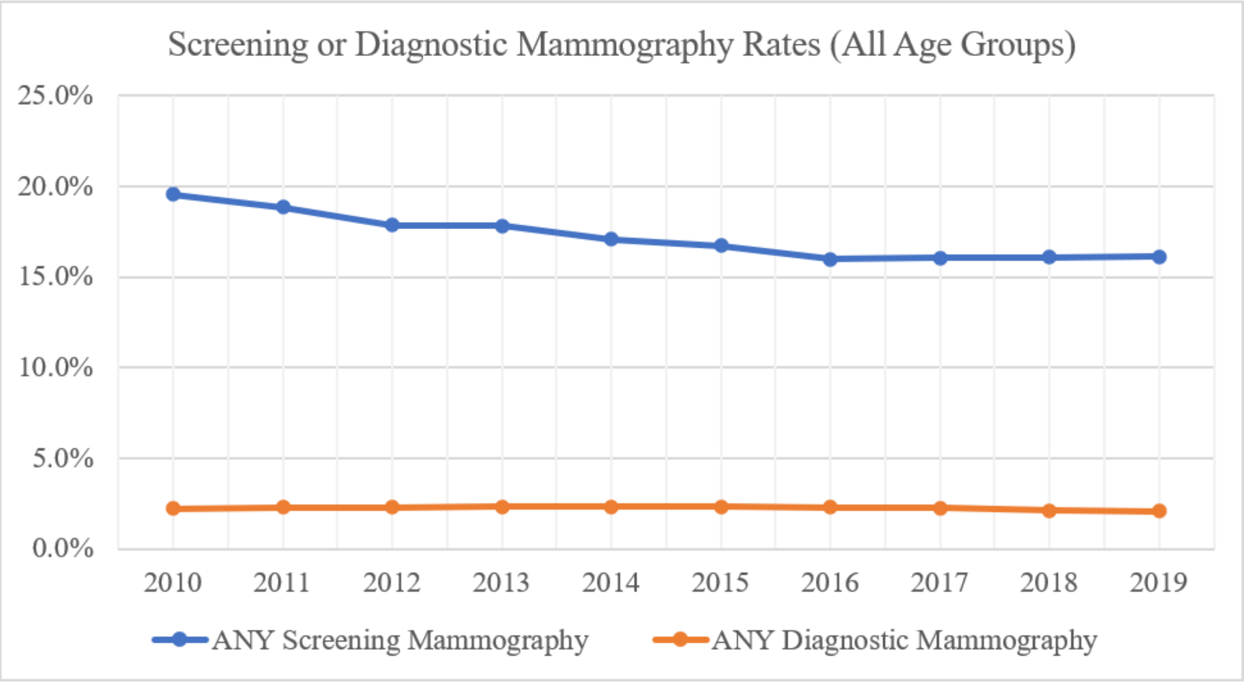

*Figure S7 Overall Screening and Diagnostic Mammography Rates, All Age Groups*

#### Colorectal Cancer Screening Figures

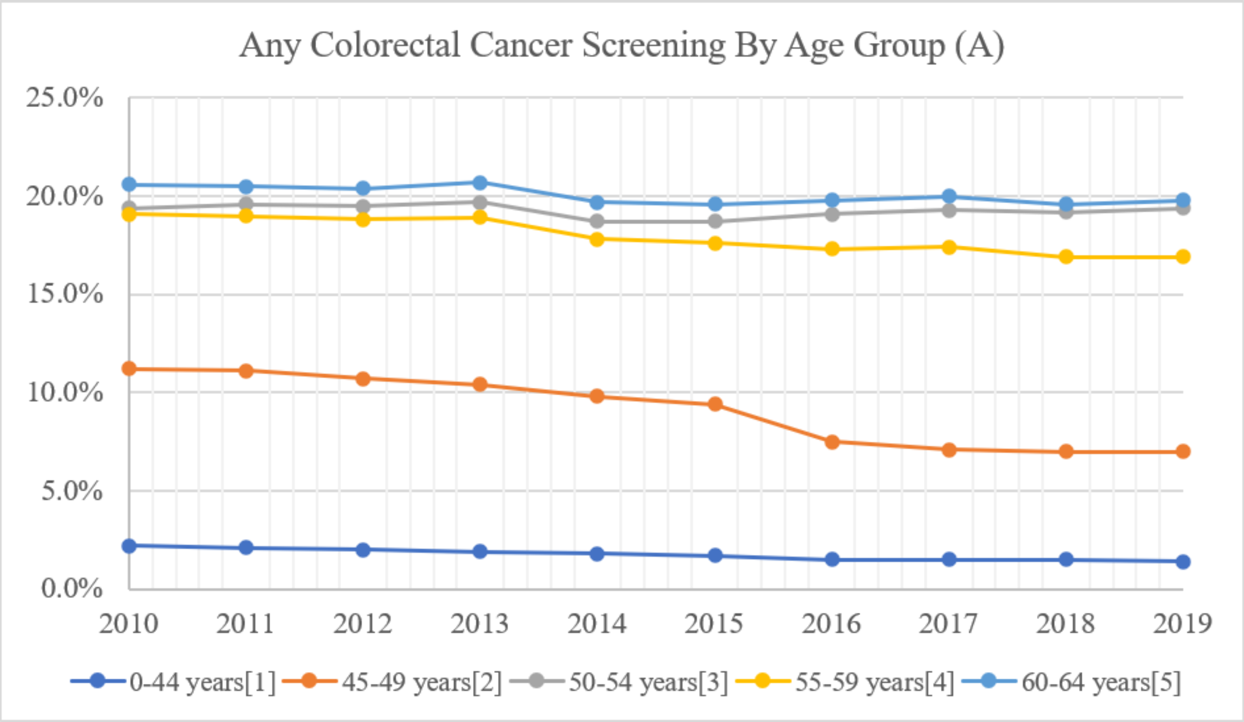

*Figure S8 Any Colorectal Cancer Screening Rate by Age Group (A)*

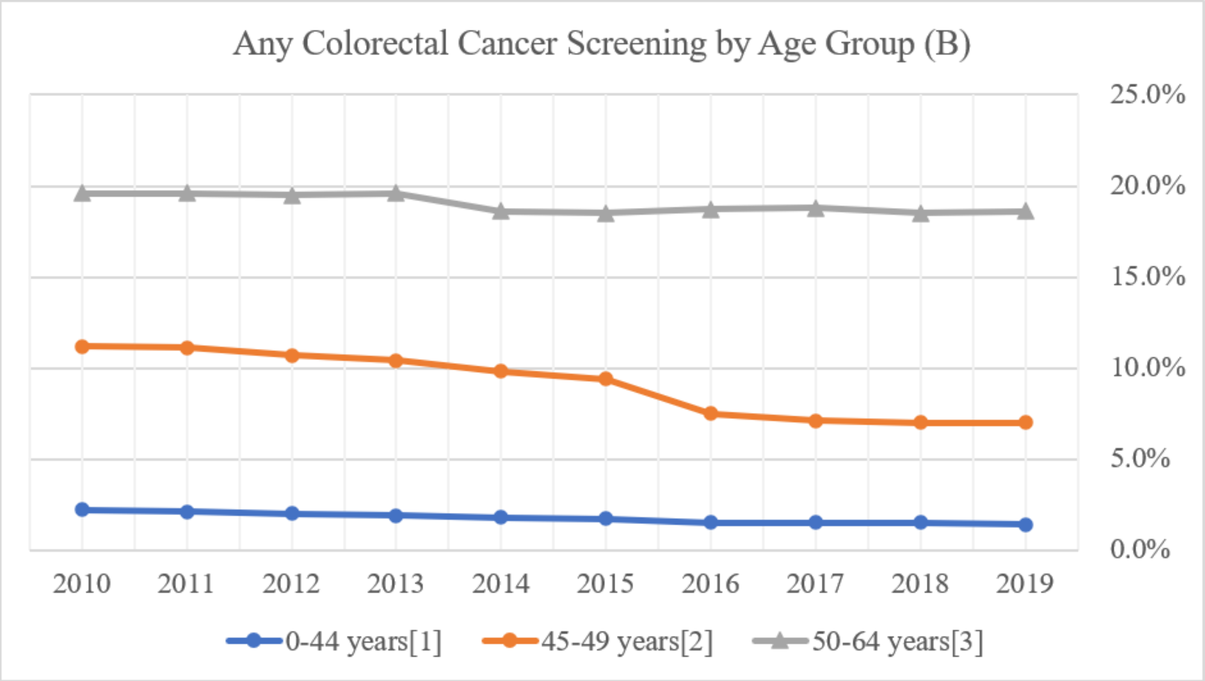

*Figure S9 Any Colorectal Cancer Screening Rate by Age Group (B)*

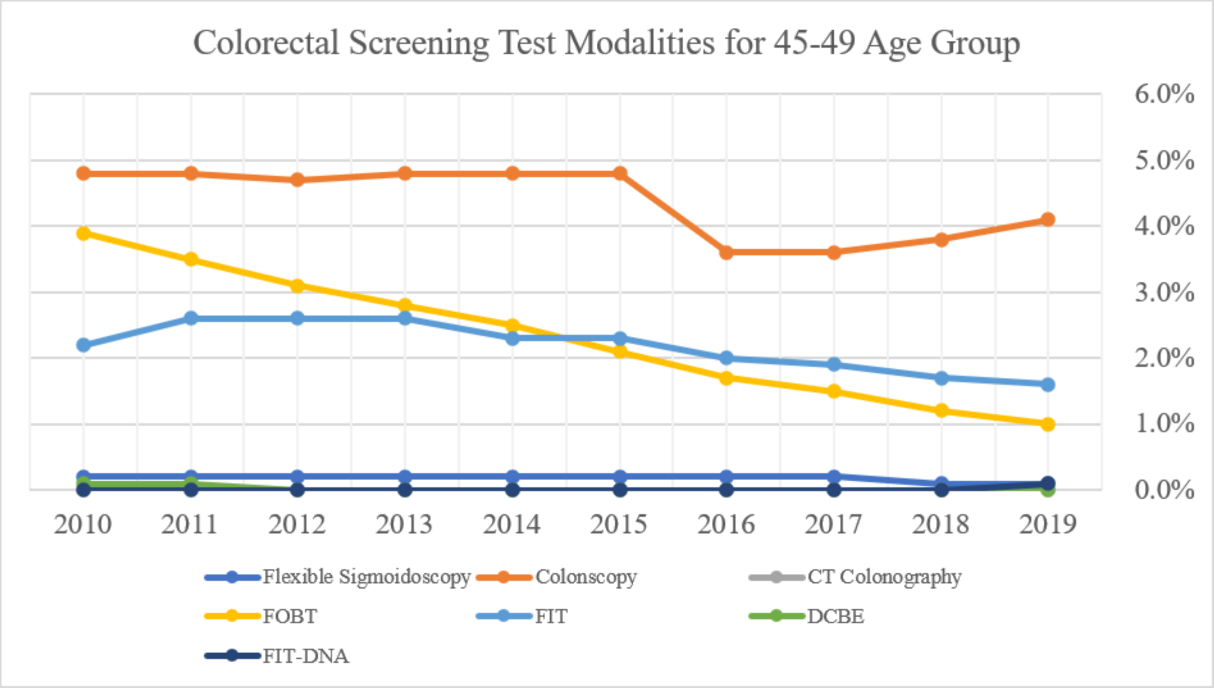

*Figure S10 Colorectal Cancer Screening by Test Modality Type, Age 45-49*

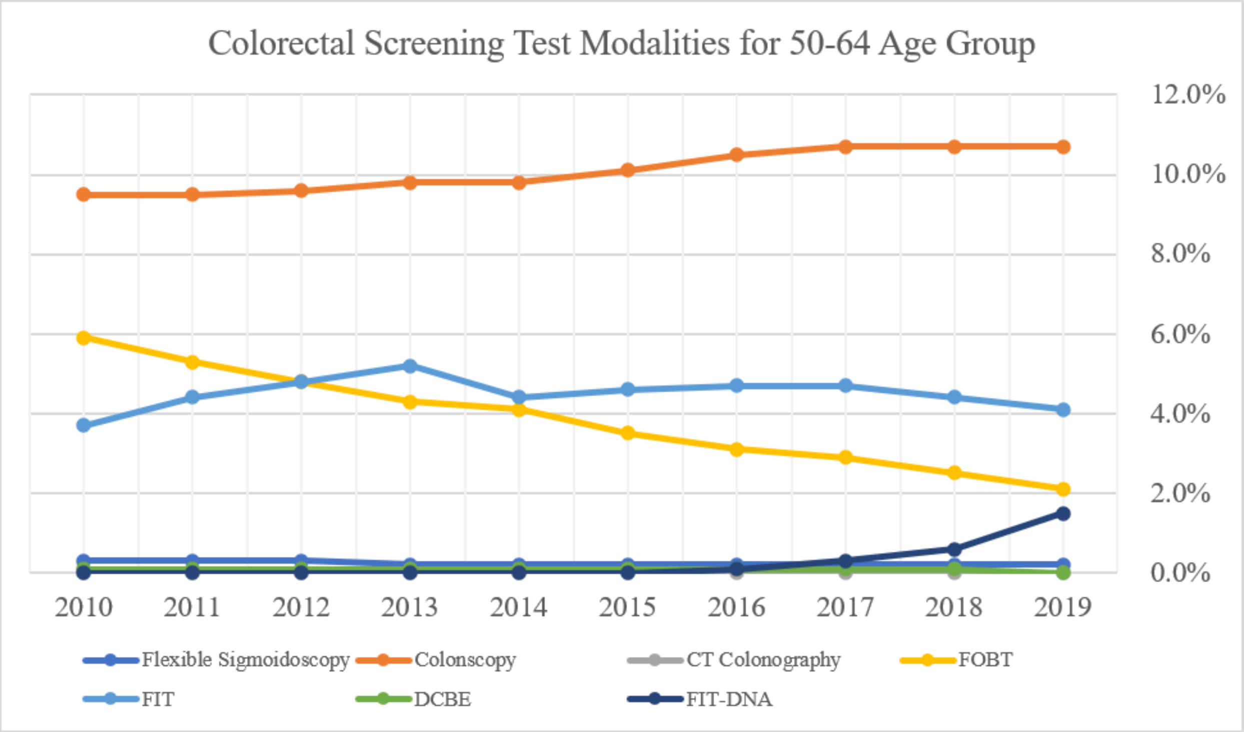

*Figure S11 Colorectal Cancer Screening by Test Modality Type, Age 50-64*

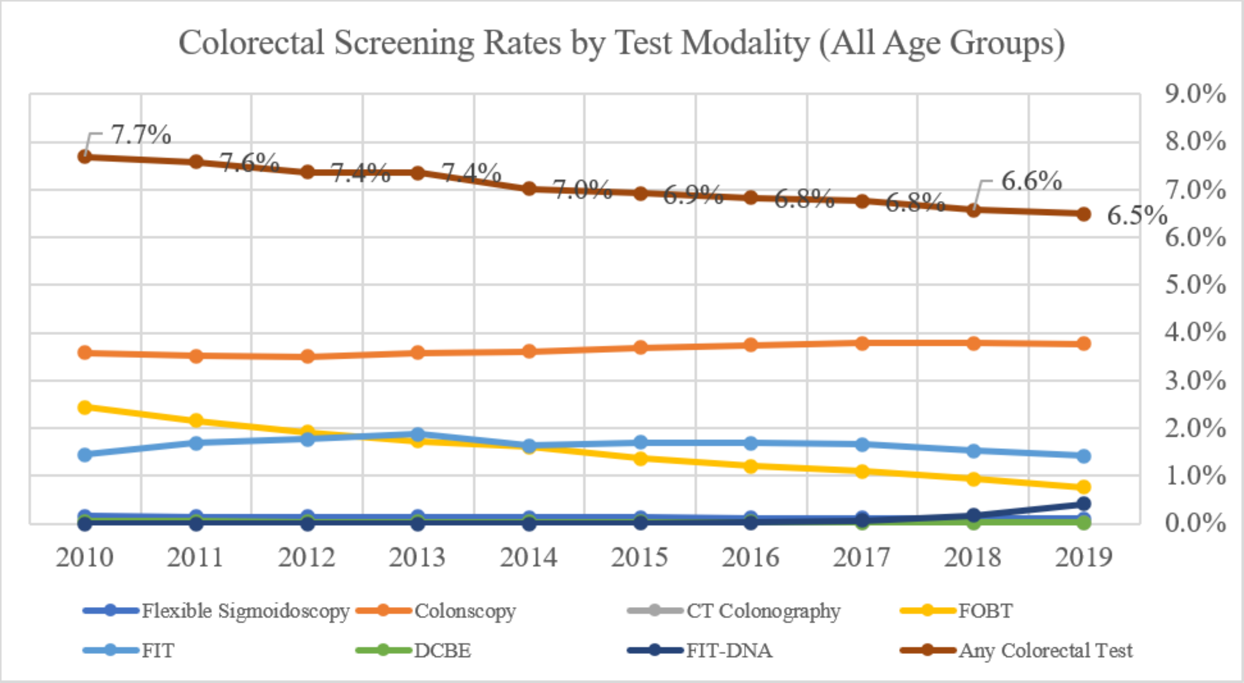

*Figure S12 Colorectal Cancer Screening Rates by Test Modalities, All Age (0-64 years)*

#### Cancer Screening Costs

*Table S1 Mean and Median Out-of-Pocket (OOP) Costs by Test Modality Type for Cervical Cancer Screening*

| | Mean (SD) OOP Cost | | | Median OOP Cost | | |
|---|---|---|---|---|---|---|
| Year | Cytology alone | HPV alone | Co-testing | Cytology alone | HPV alone | Co-testing |
| 2019 | $7.43 (34.74) | $8.12 (35.79) | $9.98 (44.58) | $0 | $0 | $0 |
| 2018 | $7.28 (35.22) | $8.47 (39.15) | $9.72 (46.22) | $0 | $0 | $0 |
| 2017 | $6.59 (33.25) | $8.02 (35.84) | $9.03 (43.25) | $0 | $0 | $0 |
| 2016 | $6.56 (31.91) | $8.42 (36.02) | $9.35 (43.11) | $0 | $0 | $0 |
| 2015 | $6.72 (31.36) | $9.19 (34.72) | $9.98 (41.26) | $0 | $0 | $0 |
| 2014 | $5.38 (28.65) | $7.37 (32.52) | $7.86 (38.44) | $0 | $0 | $0 |
| 2013 | $5.29 (26.65) | $7.52 (30.83) | $7.73 (35.78) | $0 | $0 | $0 |
| 2012 | $5.83 (25.70) | $9.99 (31.18) | $10.24 (36.67) | $0 | $0 | $0 |
| 2011 | $5.57 (23.77) | $9.62 (29.21) | $9.46 (33.94) | $0 | $0 | $0 |
| 2010 | $6.87 (24.79) | $11.20 (31.46) | $11.63 (35.93) | $0 | $0 | $0 |

Note: Out-of-pocket (OOP) cost is defined as sum of copay, coinsurance, and deductible costs.

*Table S2 Mean and Median Out-of-Pocket (OOP) Costs by Test Modality Type for Breast Cancer Screening*

| Year | Mean (SD) OOP Cost | | Median OOP Cost | |
|---|---|---|---|---|
| | ANY Screening Mammography | ANY Diagnostic Mammography | ANY Screening Mammography | ANY Diagnostic Mammography |
| 2019 | $2.98 (29.92) | $122.95 (182.45) | $0 | $35.00 |
| 2018 | $2.43 (25.86) | $106.45 (164.63) | $0 | $25.09 |
| 2017 | $2.77 (23.64) | $91.42 (143.52) | $0 | $22.11 |
| 2016 | $3.65 (27.29) | $79.78 (127.75) | $0 | $19.37 |
| 2015 | $3.28 (25.70) | $69.10 (118.67) | $0 | $12.11 |
| 2014 | $2.24 (19.79) | $54.93 (100.85) | $0 | $0 |
| 2013 | $2.59 (20.48) | $49.85 (92.18) | $0 | $0 |
| 2012 | $3.60 (24.43) | $49.65 (89.37) | $0 | $0 |
| 2011 | $4.21 (48.97) | $44.45 (81.87) | $0 | $0 |
| 2010 | $7.08 (27.90) | $39.24 (74.59) | $0 | $0 |

Note: Out-of-pocket (OOP) cost is defined as sum of copay, coinsurance, and deductible costs.

*Table S3 Mean and Median Out-of-Pocket (OOP) Costs by Test Modality Type for Colorectal Cancer Screening*

| Year | Mean (SD) OOP Cost | | | | | | | |
|---|---|---|---|---|---|---|---|---|
| | Colonoscopy | Flexible Sigmoidoscopy | CT Colonography | FIT | FOBT | FIT-DNA | DCBE | Any CRC test |
| 2019 | $130.22 (359.89) | $136.02 (311.65) | $164.12 (322.26) | $2.13 (13.75) | $1.25 (19.78) | $6.07 (52.33) | $99.14 (202.57) | $78.57 (283.57) |
| 2018 | $124.15 (352.93) | $130.83 (307.64) | $115.82 (249.97) | $2.37 (18.08) | $1.25 (16.29) | $24.47 (101.67) | $94.87 (207.05) | $74.94 (277.15) |
| 2017 | $114.59 (321.12) | $116.03 (261.08) | $112.04 (237.91) | $2.28 (13.45) | $1.24 (15.05) | $49.17 (146.51) | $86.17 (178.31) | $67.29 (248.80) |
| 2016 | $116.33 (314.79) | $106.30 (237.87) | $124.51 (251.59) | $2.45 (13.11) | $1.18 (11.88) | $118.63 (205.38) | $78.83 (164.83) | $66.95 (242.04) |
| 2015 | $109.26 (290.94) | $91.44 (215.84) | $108.18 (233.67) | $2.38 (11.28) | $1.20 (14.51) | $132.96 (209.27) | $73.69 (162.12) | $60.77 (220.63) |
| 2014 | $106.68 (316.38) | $85.10 (212.45) | $116.13 (231.08) | $2.04 (14.56) | $0.89 (12.54) | $132.96 (209.27) | $66.52 (155.55) | $57.17 (234.41) |
| 2013 | $106.19 (302.39) | $76.05 (197.84) | $104.47 (223.44) | $1.86 (14.75) | $0.87 (11.51) | $132.96 (209.27) | $61.50 (146.90) | $53.68 (218.44) |
| 2012 | $123.66 (305.59) | $76.24 (193.17) | $115.41 (239.37) | $1.92 (10.23) | $0.87 (10.75) | $0.00 (0.00) | $67.33 (162.20) | $60.72 (220.26) |
| 2011 | $131.33 (285.04) | $66.63 (163.74) | $104.82 (243.71) | $2.07 (8.94) | $0.90 (9.10) | $2.20 (0.00) | $63.57 (148.27) | $62.87 (205.43) |
| 2010 | $145.54 (279.19) | $59.12 (141.91) | $108.01 (217.92) | $2.32 (10.57) | $1.03 (9.50) | $2.20 (0.00) | $57.57 (138.19) | $69.61 (204.02) |

| Year | Median OOP Cost | | | | | | | |
|---|---|---|---|---|---|---|---|---|
| | Colonoscopy | Flexible Sigmoidoscopy | CT Colonography | FIT | FOBT | FIT-DNA | DCBE | Any CRC test |
| 2019 | $0 | $15.00 | $0 | $0 | $0 | $0 | $0 | $0 |
| 2018 | $0 | $18.84 | $0 | $0 | $0 | $0 | $0 | $0 |
| 2017 | $0 | $16.32 | $0 | $0 | $0 | $0 | $4.01 | $0 |
| 2016 | $0 | $15.75 | $0 | $0 | $0 | $0 | $0 | $0 |
| 2015 | $0 | $11.90 | $0 | $0 | $0 | $0 | $0 | $0 |
| 2014 | $0 | $8.64 | $0 | $0 | $0 | $0 | $0 | $0 |
| 2013 | $0 | $8.90 | $13.19 | $0 | $0 | $0 | $0 | $0 |
| 2012 | $0 | $10.00 | $5.00 | $0 | $0 | $0 | $0 | $0 |
| 2011 | $0 | $8.11 | $0.00 | $0 | $0 | $2.20 | $2.92 | $0 |
| 2010 | $15.97 | $13.38 | $15.00 | $0 | $0 | $2.20 | $3.46 | $0 |

Note: Out-of-pocket (OOP) cost is defined as sum of copay, coinsurance, and deductible costs.
CRC: Colorectal Cancer

### Summary of MarketScan Commercial Claims Database Population

*Table S4 Total Unique Patient Population by Claim Year, 2010-2019*

| Claim year | Overall Unique Patient Population | Total Unique Patients with 12-month continuous enrollment in a given claim year | |
|---|---|---|---|
| | Total N | N | % |
| All Years | 141,190,000 | 96,819,116 | 68.6% |
| 2019 | 22,000,413 | 15,289,626 | 69.5% |
| 2018 | 27,087,740 | 19,288,264 | 71.2% |
| 2017 | 26,146,275 | 19,248,087 | 73.6% |
| 2016 | 27,895,445 | 20,297,237 | 72.8% |
| 2015 | 28,348,363 | 20,433,783 | 72.1% |
| 2014 | 47,258,528 | 32,685,899 | 69.2% |
| 2013 | 43,737,217 | 30,811,938 | 70.4% |
| 2012 | 53,131,420 | 37,447,218 | 70.5% |
| 2011 | 53,012,885 | 37,532,147 | 70.8% |
| 2010 | 51,672,523 | 36,740,536 | 71.1% |
| Annual Summary | | | |
| Average | 38,029,081 | 26,977,474 | 71.1% |
| Median | 36,042,790 | 25,622,861 | 71.0% |
| Min | 22,000,413 | 15,289,626 | 69.2% |
| Max | 53,131,420 | 37,532,147 | 73.6% |

### Additional Results

*Table S5 2019 Unadjusted Cancer Screening Rates by Place of Residence*

| Place of Residence | Cervical Cancer Screening in 2019 | | | Breast Cancer Screening in 2019 | | | Colorectal Cancer Screening in 2019 | | |
|---|---|---|---|---|---|---|---|---|---|
| | Rate | n (%) | Risk Difference (95% CI) | Rate | n (%) | Risk Difference (95% CI) | Rate | n (%) | Risk Difference (95% CI) |
| Rural | 26.47% | 566,500 (10.91%) | -4.17% (-4.29% to -4.04%) | 41.48% | 288,158 (12.15%) | -1.20% (-1.39% to -1.00%) | 14.22% | 658,245 (12.06%) | -1.88% (-1.97% to -1.79%) |
| Urban | 30.63% | 4,113,538 (79.23%) | Ref | 42.67% | 1,846,287 (77.86%) | Ref | 16.10% | 4,275,838 (78.33%) | Ref |
| Unknown | 28.12% | 512,065 (9.86%) | -2.51% (-2.64% to -2.38%) | 41.26% | 236,933 (9.99%) | -1.42% (-1.63% to -1.21%) | 14.51% | 524,578 (9.61%) | -1.59% (-1.69% to -1.49%) |
| Total | 29.93% | 5,192,103 (100.00%) | | 42.39% | 2,371,378 (100.00%) | | 15.72% | 5,458,661 (100.00%) | |

### List of Procedure and Diagnosis Codes

#### Cervical Cancer Screening

##### *Cervical Cancer Screening Inclusion Codes*

*Table S6 List of Procedure and Diagnosis Codes for Cervical Cancer Screening, including Cytology and HPV Testing*

| Cytology codes | | |
|---|---|---|
| Type | Code | Code Description |
| CPT | 88141 | Cytopathology, cervical or vaginal (any reporting system), requiring interpretation by physician |
| CPT | 88142 | Cytopathology, cervical or vaginal (any reporting system), collected in preservative fluid, automated thin layer preparation; manual screening under physician supervision |
| CPT | 88143 | Cytopathology, cervical or vaginal (any reporting system), collected in preservative fluid, automated thin layer preparation; with manual screening and rescreening under physician supervision |
| CPT | 88147 | Cytopathology smears, cervical or vaginal; screening by automated system under physician supervision |
| CPT | 88148 | Cytopathology smears, cervical or vaginal; screening by automated system with manual rescreening under physician supervision |
| CPT | 88150 | Cytopathology, slides, cervical or vaginal; manual screening under physician supervision |
| CPT | 88152 | Cytopathology, slides, cervical or vaginal; with manual screening and computer-assisted rescreening under physician supervision |
| CPT | 88153 | Cytopathology, slides, cervical or vaginal; with manual screening and rescreening under physician supervision |
| CPT | 88154 | Cytopathology, slides, cervical or vaginal; with manual screening and computer-assisted rescreening using cell selection and review under physician supervision |
| CPT | 88155 | C ytopathology, slides, cervical or vaginal, definitive hormonal evaluation (e.g., maturation index, karyopyknotic index, estrogenic index) (List separately in addition to code[s] for other technical and interpretation services) |
| CPT | 88164 | Cytopathology, slides, cervical or vaginal (the Bethesda System); manual screening under physician supervision |
| CPT | 88165 | Cytopathology, slides, cervical or vaginal (the Bethesda System); with manual screening and rescreening under physician supervision |
| CPT | 88166 | Cytopathology, slides, cervical or vaginal (the Bethesda System); with manual screening and computer-assisted rescreening under physician supervision |

| | | |
|---|---|---|
| CPT | 88167 | Cytopathology, slides, cervical or vaginal (the Bethesda System); with manual screening and computer-assisted rescreening using cell selection and review under physician supervision |
| CPT | 88174 | Cytopathology, cervical or vaginal (any reporting system), collected in preservative fluid, automated thin layer preparation; screening by automated system, under physician supervision |
| CPT | 88175 | Cytopathology, cervical or vaginal (any reporting system), collected in preservative fluid, automated thin layer preparation; with screening by automated system and manual rescreening or review, under physician supervision |
| HCPCS | G0101 | Cervical or vaginal cancer screening; pelvic and clinical breast examination |
| HCPCS | G0123 | Screening cytopathology, cervical or vaginal (any reporting system, collected in preservative fluid, automated thin layer preparation, screening by cytotechnologist under physician supervision |
| HCPCS | G0124 | Screening cytopathology, cervical or vaginal (any reporting system), collected in preservative fluid, automated thin layer preparation, requiring interpretation by physician |
| HCPCS | G0141 | Screening cytopathology smears, cervical or vaginal, performed by automated system, with manual rescreening, requiring interpretation by physician |
| HCPCS | G0143 | Screening cytopathology, cervical or vaginal (any reporting system), collected in preservative fluid, automated thin layer preparation, with manual screening and rescreening by cytotechnologist under physician supervision |
| HCPCS | G0144 | Screening cytopathology, cervical or vaginal (any reporting system), collected in preservative fluid, automated thin layer preparation, with screening by automated system, under physician supervision |
| HCPCS | G0145 | Screening cytopathology, cervical or vaginal (any reporting system), collected in preservative fluid, automated thin layer preparation, with screening by automated system and manual rescreening under physician supervision |
| HCPCS | G0147 | Screening cytopathology smears, cervical or vaginal, performed by automated system under physician supervision |
| HCPCS | G0148 | Screening cytopathology smears, cervical or vaginal, performed by automated system with manual rescreening |
| HCPCS | P3000 | Screening Papanicolaou smear, cervical or vaginal, up to three smears, by technician under physician supervision |
| HCPCS | P3001 | Screening Papanicolaou smear, cervical or vaginal, up to three smears, requiring interpretation by physician |
| HCPCS | Q0091 | Screening Papanicolaou smear; obtaining, preparing and conveyance of cervical or vaginal smear to laboratory |
| ICD-9-CM | 91.5 | Microscopic examination of specimen from female genital tract, cell block and Papanicolaou smear |
| ICD-9-CM | V76.2 | Routine cervical Papanicolaou smear |

| | | |
|---|---|---|
| ICD-10-CM | Z12.4 | Encounter for screening pap smear for malignant neoplasm of cervix |
| HPV testing | | |
| Type | Code | Code Description |
| CPT | 0500T | Infectious agent detection by nucleic acid (DNA or RNA), human papillomavirus (HPV) for five or more separately reported high-risk HPV types (eg, 16, 18, 31, 33, 35, 39, 45, 51, 52, 56, 58, 59, 68) (i.e., genotyping) |
| CPT | 87620 | Infectious agent detection by nucleic acid (DNA or RNA); papillomavirus, human, direct probe technique |
| CPT | 87621 | Infectious agent detection by nucleic acid (DNA or RNA); papillomavirus, human, amplified probe technique |
| CPT | 87622 | Infectious agent detection by nucleic acid (DNA or RNA); papillomavirus, human, quantification |
| CPT | 87624 | Infectious agent detection by nucleic acid (DNA or RNA); HPV, high-risk types (e.g., 16, 18, 31, 33, 35, 39, 45, 51, 52, 56, 58, 59, 68) |
| CPT | 87625 | Infectious agent detection by nucleic acid (DNA or RNA); HPV, types 16 and 18 only, includes type 45, if performed |
| HCPCS | G0476 | Infectious agent detection by nucleic acid (DNA or RNA); HPV, high-risk types (e.g., 16, 18, 31, 33, 35, 39, 45, 51, 52, 56, 58, 59, 68) for cervical cancer screening, must be performed in addition to pap test |
| ICD-9-CM | V73.81 | Special screening examination for Human papillomavirus (HPV) |
| ICD-10-CM | Z11.51 | Encounter for screening for human papillomavirus (HPV) |

Source: (1-6)

##### *Cervical Cancer Exclusion Codes*

*Table S7 List of Exclusionary Procedure and Diagnosis Codes for Cervical Cancer Screening, including Hysterectomy and Evidence of Prior Cervical Cancer Diagnosis*

| Exclusion Codes | | |
|---|---|---|
| Type | Code | Description |
| CPT | 51925 | Closure of vesicouterine fistula; with hysterectomy |
| CPT | 56308 | laparoscopy, surgical; with vaginal hysterectomy with or without removal of tube[s], with or without removal of ovary[s] [laproscopic assisted vaginal hysterectomy] |
| CPT | 57540 | Excision of cervical stump, abdominal approach |
| CPT | 57545 | Excision of cervical stump, abdominal approach; with pelvic floor repair |
| CPT | 57550 | Excision of cervical stump, vaginal approach |
| CPT | 57555 | Excision of cervical stump, vaginal approach; with anterior and/or posterior repair |
| CPT | 57556 | Excision of cervical stump, vaginal approach; with repair of enterocele |

| | | |
|---|---|---|
| CPT | 58150 | Total abdominal hysterectomy (corpus and cervix), with or without removal of tube(s), with or without removal of ovary(s) |
| CPT | 58152 | Total abdominal hysterectomy (corpus and cervix), with or without removal of tube(s), with or without removal of ovary(s) |
| CPT | 58200 | Total abdominal hysterectomy, including partial vaginectomy, with para-aortic and pelvic lymph node sampling, with or without removal of tube(s), with or without removal of ovary(s) |
| CPT | 58210 | Radical abdominal hysterectomy, with bilateral total pelvic lymphadenectomy and para-aortic lymph node sampling (biopsy), with or without removal of tube(s), with or without removal of ovary(s) |
| CPT | 58240 | Pelvic exenteration for gynecologic malignancy, with total abdominal hysterectomy or cervicectomy, with or without removal of tube(s), with or without removal of ovary(s), with removal of bladder and ureteral transplantations, and/or abdominoperineal resection of rectum and colon and colostomy, or any combination thereof |
| CPT | 58260 | Vaginal hysterectomy, for uterus 250 g or less |
| CPT | 58262 | Vaginal hysterectomy, for uterus 250 g or less; with removal of tube(s), and/or ovary(s) |
| CPT | 58263 | Vaginal hysterectomy, for uterus 250 g or less; with removal of tube(s), and/or ovary(s), with repair of enterocele |
| CPT | 58267 | Vaginal hysterectomy, for uterus 250 g or less; with colpo-urethrocystopexy (Marshall-Marchetti-Krantz type, Pereyra type) with or without endoscopic control |
| CPT | 58270 | Vaginal hysterectomy, for uterus 250 g or less; with repair of enterocele |
| CPT | 58275 | Vaginal hysterectomy, with total or partial vaginectomy |
| CPT | 58280 | Vaginal hysterectomy, with total or partial vaginectomy; with repair of enterocele |
| CPT | 58285 | Vaginal hysterectomy, radical (Schauta type operation) |
| CPT | 58290 | Vaginal hysterectomy, for uterus greater than 250 g |
| CPT | 58291 | Vaginal hysterectomy, for uterus greater than 250 g; with removal of tube(s) and/or ovary(s) |
| CPT | 58292 | Vaginal hysterectomy, for uterus greater than 250 g; with removal of tube(s) and/or ovary(s), with repair of enterocele |
| CPT | 58293 | Vaginal hysterectomy, for uterus greater than 250 g; with colpo-urethrocystopexy (Marshall-Marchetti-Krantz type, Pereyra type) with or without endoscopic control |
| CPT | 58294 | Vaginal hysterectomy, for uterus greater than 250 g; with repair of enterocele |
| CPT | 56308 | Laparoscopy, surgical; with vaginal hysterectomy with or without removal of tube[s], with or without removal of ovary[s] [laproscopic assisted vaginal hysterectomy] |
| CPT | 58548 | Laparoscopy, surgical, with radical hysterectomy, with bilateral total pelvic lymphadenectomy and para-aortic lymph node sampling (biopsy), with removal of tube(s) and ovary(s), if performed |
| CPT | 58550 | Laparoscopy, surgical, with vaginal hysterectomy, for uterus 250 g or less |

| | | |
|---|---|---|
| CPT | 58552 | Laparoscopy, surgical, with vaginal hysterectomy, for uterus 250 g or less; with removal of tube(s) and/or ovary(s) |
| CPT | 58553 | Laparoscopy, surgical, with vaginal hysterectomy, for uterus greater than 250 g |
| CPT | 58554 | Laparoscopy, surgical, with vaginal hysterectomy, for uterus greater than 250 g; with removal of tube(s) and/or ovary(s) |
| CPT | 58570 | Laparoscopy, surgical, with total hysterectomy, for uterus 250 g or less |
| CPT | 58571 | Laparoscopy, surgical, with total hysterectomy, for uterus 250 g or less; with removal of tube(s) and/or ovary(s) |
| CPT | 58572 | Laparoscopy, surgical, with total hysterectomy, for uterus greater than 250 g |
| CPT | 58573 | Laparoscopy, surgical, with total hysterectomy, for uterus greater than 250 g; with removal of tube(s) and/or ovary(s) |
| CPT | 58951 | Resection (initial) of ovarian, tubal or primary peritoneal malignancy with bilateral salpingo-oophorectomy and omentectomy; with total abdominal hysterectomy, pelvic and limited para-aortic lymphadenectomy |
| CPT | 59525 | Subtotal or total hysterectomy after cesarean delivery (List separately in addition to code for primary procedure) |
| CPT | 58953 | Bilateral salpingo-oophorectomy with omentectomy, total abdominal hysterectomy and radical dissection for debulking |
| CPT | 58954 | Bilateral salpingo-oophorectomy with omentectomy, total abdominal hysterectomy and radical dissection for debulking; with pelvic lymphadenectomy and limited para-aortic lymphadenectomy |
| CPT | 58956 | Bilateral salpingo-oophorectomy with total omentectomy, total abdominal hysterectomy for malignancy |
| CPT | 59135 | Surgical treatment of ectopic pregnancy; interstitial, uterine pregnancy requiring total hysterectomy |
| CPT | 58575 | Laparoscopy, surgical, total hysterectomy for resection of malignancy (tumor debulking), with omentectomy including salpingo-oophorectomy, unilateral or bilateral, when performed |
| CPT | 59525 | Subtotal or total hysterectomy after cesarean delivery (List separately in addition to code for primary procedure) |
| CPT | 57522 | Conization of cervix, with or without fulguration, with or without dilation and curettage, with or without repair; loop electrode excision |
| CPT | 57460 | Colposcopy of the cervix including upper/adjacent vagina; with loop electrode biopsy(s) of the cervix |
| CPT | 57461 | Colposcopy of the cervix including upper/adjacent vagina; with loop electrode conization of the cervix |
| CPT | 57510 | Cautery of cervix; electro or thermal |
| CPT | 57511 | Cautery of cervix; cryocautery, initial or repeat |
| CPT | 57510 | Cautery of cervix; electro or thermal |
| CPT | 57513 | Cautery of cervix; laser ablation |
| CPT | 57420 | Colposcopy of the entire vagina, with cervix if present |
| CPT | 57421 | Colposcopy of the entire vagina, with cervix if present; with biopsy(s) of vagina/cervix |

| | | |
|---|---|---|
| CPT | 57505 | Endocervical curettage (not done as part of a dilation and curettage) |
| CPT | 57500 | Biopsy of cervix, single or multiple, or local excision of lesion, with or without fulguration (separate procedure) |
| CPT | 57452 | Colposcopy of the cervix including upper/adjacent vagina |
| CPT | 57454 | Colposcopy of the cervix including upper/adjacent vagina with biopsy(s) of the cervix and endocervical curettage |
| CPT | 57455 | Colposcopy of the cervix including upper/adjacent vagina with biopsy(s) of the cervix |
| CPT | 57456 | Colposcopy of the cervix including upper/adjacent vagina with endocervical curettage |
| CPT | 57460 | Colposcopy of the cervix including upper/adjacent vagina with loop electrode biopsy(s) of the cervix |
| CPT | 57461 | Colposcopy of the cervix including upper/adjacent vagina with loop electrode conization of the cervix |
| CPT | 57520 | Conization of cervix, with or without fulguration, with or without dilation and curettage, with or without repair; cold knife or laser |
| ICD-10-CM | C53 | Malignant neoplasm of cervix uteri |
| ICD-10-CM | C53.0 | Malignant neoplasm of endocervix |
| ICD-10-CM | C53.1 | Malignant neoplasm of exocervix |
| ICD-10-CM | C53.8 | Malignant neoplasm of overlapping sites of cervix uter |
| ICD-10-CM | C53.9 | Malignant neoplasm of cervix uteri, unspecified |
| ICD-10-CM | Z85.41 | Personal history of malignant neoplasm of cervix uteri |
| ICD-10-CM | Z87.410 | Personal history of cervical dysplasia |
| ICD-10-CM | Z90.710 | Acquired absence of both cervix and uterus |
| ICD-10-CM | Z90.712 | Acquired absence of cervix with remaining uterus |
| ICD-10-CM | Z12.72 | Encounter for screening for malignant neoplasm of vagina |
| ICD-10-PCS | 0UTC0ZZ | Resection of Cervix, Open Approach |
| ICD-10-PCS | 0UTC4ZZ | Resection of Cervix, Percutaneous Endoscopic Approach |
| ICD-10-PCS | 0UTC7ZZ | Resection of Cervix, Via Natural or Artificial Opening |
| ICD-10-PCS | 0UTC8ZZ | Resection of Cervix, Via Natural or Artificial Opening Endoscopic |
| ICD-10-PCS | 0UT90ZZ | Resection of Uterus, Open Approach |

| | | |
|---|---|---|
| ICD-10-PCS | 0UT94ZZ | Resection of Uterus, Percutaneous Endoscopic Approach |
| ICD-10-PCS | 0UT97ZZ | Resection of Uterus, Via Natural or Artificial Opening |
| ICD-10-PCS | 0UT98ZZ | Resection of Uterus, Via Natural or Artificial Opening Endoscopic |
| ICD-10-PCS | 0UT9FZZ | Resection of Uterus, Via Natural or Artificial Opening With Percutaneous Endoscopic Assistance |
| ICD-10-PCS | 0U5C0ZZ | Destruction of Cervix, Open Approach |
| ICD-10-PCS | 0U5C3ZZ | Destruction of Cervix, Percutaneous Approach |
| ICD-10-PCS | 0U5C4ZZ | Destruction of Cervix, Percutaneous Endoscopic Approach |
| ICD-10-PCS | 0U5C7ZZ | Destruction of Cervix, Via Natural or Artificial Opening |
| ICD-10-PCS | 0U5C8ZZ | Destruction of Cervix, Via Natural or Artificial Opening Endoscopic |
| ICD-10-PCS | 0UBC0ZX | Excision of Cervix, Open Approach, Diagnostic |
| ICD-10-PCS | 0UBC3ZX | Excision of Cervix, Percutaneous Approach, Diagnostic |
| ICD-10-PCS | 0UBC4ZX | Excision of Cervix, Percutaneous Endoscopic Approach, Diagnostic |
| ICD-10-PCS | 0UBC7ZX | Excision of Cervix, Via Natural or Artificial Opening, Diagnostic |
| ICD-10-PCS | 0UBC8ZX | Excision of Cervix, Via Natural or Artificial Opening Endoscopic, Diagnostic |
| ICD-9-CM | 67.32 | Destruction of lesion of cervix by cauterization |
| ICD-9-CM | 67.33 | Destruction of lesion of cervix by cryosurgery |
| ICD-9-CM | 180 | Malignant neoplasm of cervix uteri |
| ICD-9-CM | 180 | Malignant neoplasm of endocervix |
| ICD-9-CM | 180.1 | Malignant neoplasm of exocervix |
| ICD-9-CM | 180.8 | Malignant neoplasm of other specified sites of cervix |
| ICD-9-CM | 180.9 | Malignant neoplasm of cervix uteri, unspecified site |
| ICD-9-CM | V10.41 | Personal history of malignant neoplasm of cervix uteri |
| ICD-9-CM | V13.22 | Personal history of cervical dysplasia |
| ICD-9-CM | V72.32 | Encounter for Papanicolaou cervical smear to confirm findings of recent normal smear following initial abnormal smear |
| ICD-9-CM | V76.47 | Special screening for malignant neoplasms of vagina |
| ICD-9-CM | V88.01 | Acquired absence of both cervix and uterus |
| ICD-9-CM | V88.03 | Acquired absence of cervix with remaining uterus |
| ICD-9-PCS | 67.11 | Endocervical biopsy |

| | | |
|---|---|---|
| ICD-9-PCS | 67.12 | Other cervical biopsy |
| ICD-9-PCS | 67.19 | Other diagnostic procedures on cervix |
| ICD-9-PCS | 67.2 | Conization of cervix |
| ICD-9-PCS | 68.41 | Laparoscopic total abdominal hysterectomy |
| ICD-9-PCS | 68.49 | Other and unspecified total abdominal hysterectomy |
| ICD-9-PCS | 68.51 | Laparoscopically assisted vaginal hysterectomy (LAVH) |
| ICD-9-PCS | 68.59 | Other and unspecified vaginal hysterectomy |
| ICD-9-PCS | 68.61 | Laparoscopic radical abdominal hysterectomy |
| ICD-9-PCS | 68.69 | Other and unspecified radical abdominal hysterectomy |
| ICD-9-PCS | 68.71 | Laparoscopic radical vaginal hysterectomy [LRVH] |
| ICD-9-PCS | 68.79 | Other and unspecified radical vaginal hysterectomy |
| ICD-9-PCS | 68.8 | Pelvic evisceration |
| ICD-9-PCS | 68.9 | Other and unspecified hysterectomy |

Source: (1-6)

#### Breast Cancer Screening

##### *Breast Cancer Screening Inclusion Codes*

*Table S8 List of Procedure Codes for Breast Cancer Screening, including Screening and Diagnostic Mammography*

| Screening Mammography | | |
|---|---|---|
| Type | Code | Description |
| CPT | 77052 | Computer-aided detection (computer algorithm analysis of digital image data for lesion detection) with further review for interpretation, with or without digitization of film radiographic images; screening mammography (List separately in addition to code for primary procedure) |
| CPT | 77057 | Screening mammography, bilateral (2-view study of each breast) |
| HCPCS | G0202 | Screening mammography, bilateral (2-view study of each breast), including computer-aided detection (cad) when performed |

| | | |
|---|---|---|
| CPT | 77067 | Screening mammography, bilateral (2-view study of each breast), including computer-aided detection (CAD) when performed |
| CPT | 77063 | Screening digital breast tomosynthesis, bilateral (List separately in addition to code for primary procedure) |
| Diagnostic Mammography | | |
| Type | Code | Description |
| CPT | 77051 | Computer-aided detection (computer algorithm analysis of digital image data for lesion detection) with further review for interpretation, with or without digitization of film radiographic images; diagnostic mammography (list separately in addition to code for primary procedure) |
| CPT | 77055 | Mammography; unilateral |
| CPT | 77056 | Mammography; bilateral |
| HCPCS | G0204 | Diagnostic mammography, including computer-aided detection (cad) when performed; bilateral |
| HCPCS | G0206 | Diagnostic mammography, including computer-aided detection (cad) when performed; unilateral |
| CPT | 77065 | Diagnostic mammography, including computer-aided detection (CAD) when performed; unilateral |
| CPT | 77066 | Diagnostic mammography, including computer-aided detection (CAD) when performed; bilateral |
| HCPCS | G0279 | Diagnostic digital breast tomosynthesis, unilateral or bilateral (list separately in addition to 77065 or 77066) |
| CPT | 77061 | Diagnostic digital breast tomosynthesis; unilateral |
| CPT | 77062 | Diagnostic digital breast tomosynthesis; bilateral |
| ICD-9-PCS | 87.36 | Xerography of breast |
| ICD-9-PCS | 87.37 | Other mammography |

Note: Only CPT and HCPCS codes effective during the study period (Jan 1, 2010 thru Dec 31, 2019) were included for analysis. Other codes for screening (e.g., G0203,76085,76083,76092) or diagnostic purposes (e.g., G0205,G0207,76090,76091,76082) were not included.

Sources: (1-5, 7-9)

##### *Breast Cancer Screening Exclusion Codes*

*Table S9 List of Exclusionary Procedure and Diagnosis Codes for Breast Cancer, including Mastectomy and Evidence of Prior Breast Cancer Diagnosis*

| Breast cancer diagnosis codes | | |
|---|---|---|
| Type | Code | Descriptor |
| ICD-10-CM | C50.011 | Malignant neoplasm of nipple and areola, right female breast |

| | | |
|---|---|---|
| ICD-10-CM | C50.012 | Malignant neoplasm of nipple and areola, left female breast |
| ICD-10-CM | C50.019 | Malignant neoplasm of nipple and areola, unspecified female breast |
| ICD-10-CM | C50.111 | Malignant neoplasm of central portion of right female breast |
| ICD-10-CM | C50.112 | Malignant neoplasm of central portion of left female breast |
| ICD-10-CM | C50.119 | Malignant neoplasm of central portion of unspecified female breast |
| ICD-10-CM | C50.211 | Malignant neoplasm of upper-inner quadrant of right female breast |
| ICD-10-CM | C50.212 | Malignant neoplasm of upper-inner quadrant of left female breast |
| ICD-10-CM | C50.219 | Malignant neoplasm of upper-inner quadrant of unspecified female breast |
| ICD-10-CM | C50.311 | Malignant neoplasm of lower-inner quadrant of right female breast |
| ICD-10-CM | C50.312 | Malignant neoplasm of lower-inner quadrant of left female breast |
| ICD-10-CM | C50.319 | Malignant neoplasm of lower-inner quadrant of unspecified female breast |
| ICD-10-CM | C50.411 | Malignant neoplasm of upper-outer quadrant of right female breast |
| ICD-10-CM | C50.412 | Malignant neoplasm of upper-outer quadrant of left female breast |
| ICD-10-CM | C50.419 | Malignant neoplasm of upper-outer quadrant of unspecified female breast |
| ICD-10-CM | C50.511 | Malignant neoplasm of lower-outer quadrant of right female breast |
| ICD-10-CM | C50.512 | Malignant neoplasm of lower-outer quadrant of left female breast |
| ICD-10-CM | C50.519 | Malignant neoplasm of lower-outer quadrant of unspecified female breast |
| ICD-10-CM | C50.611 | Malignant neoplasm of axillary tail of right female breast |
| ICD-10-CM | C50.612 | Malignant neoplasm of axillary tail of left female breast |
| ICD-10-CM | C50.619 | Malignant neoplasm of axillary tail of unspecified female breast |
| ICD-10-CM | C50.811 | Malignant neoplasm of overlapping sites of right female breast |
| ICD-10-CM | C50.812 | Malignant neoplasm of overlapping sites of left female breast |
| ICD-10-CM | C50.819 | Malignant neoplasm of overlapping sites of unspecified female breast |
| ICD-10-CM | C50.911 | Malignant neoplasm of unspecified site of right female breast |
| ICD-10-CM | C50.912 | Malignant neoplasm of unspecified site of left female breast |
| ICD-10-CM | C50.919 | Malignant neoplasm of unspecified site of unspecified female breast |
| ICD-9-CM | 174.0 | Malignant neoplasm of nipple and areola of female breast |
| ICD-9-CM | 174.1 | Malignant neoplasm of central portion of female breast |
| ICD-9-CM | 174.2 | Malignant neoplasm of upper-inner quadrant of female breast |
| ICD-9-CM | 174.3 | Malignant neoplasm of lower-inner quadrant of female breast |
| ICD-9-CM | 174.4 | Malignant neoplasm of upper-outer quadrant of female breast |
| ICD-9-CM | 174.5 | Malignant neoplasm of lower-outer quadrant of female breast |
| ICD-9-CM | 174.6 | Malignant neoplasm of axillary tail of female breast |
| ICD-9-CM | 174.8 | Malignant neoplasm of other specified sites of female breast |
| ICD-9-CM | 174.9 | Malignant neoplasm of breast (female), unspecified |
| ICD-10-CM | C79.2 | Secondary malignant neoplasm of skin |
| ICD-10-CM | C79.81 | Secondary malignant neoplasm of breast |
| ICD-10-CM | D05.00 | Lobular carcinoma in situ of unspecified breast |
| ICD-10-CM | D05.01 | Lobular carcinoma in situ of right breast |
| ICD-10-CM | D05.02 | Lobular carcinoma in situ of left breast |
| ICD-10-CM | D05.10 | Intraductal carcinoma in situ of unspecified breast |
| ICD-10-CM | D05.11 | Intraductal carcinoma in situ of right breast |
| ICD-10-CM | D05.12 | Intraductal carcinoma in situ of left breast |

| | | |
|---|---|---|
| ICD-10-CM | D05.80 | Other specified type of carcinoma in situ of unspecified breast |
| ICD-10-CM | D05.81 | Other specified type of carcinoma in situ of right breast |
| ICD-10-CM | D05.82 | Other specified type of carcinoma in situ of left breast |
| ICD-10-CM | D05.90 | Unspecified type of carcinoma in situ of unspecified breast |
| ICD-10-CM | D05.91 | Unspecified type of carcinoma in situ of right breast |
| ICD-10-CM | D05.92 | Unspecified type of carcinoma in situ of left breast |
| ICD-10-CM | D48.60 | Neoplasm of uncertain behavior of unspecified breast |
| ICD-10-CM | D48.61 | Neoplasm of uncertain behavior of right breast |
| ICD-10-CM | D48.62 | Neoplasm of uncertain behavior of left breast |
| ICD-10-CM | D49.3 | Neoplasm of unspecified behavior of breast |
| ICD-10-CM | Z85.3 | Personal history of malignant neoplasm of breast |
| ICD-9-CM | 198.2 | Secondary malignant neoplasm of skin |
| ICD-9-CM | 198.81 | Secondary malignant neoplasm of breast |
| ICD-9-CM | 233.0 | Carcinoma in situ of breast |
| ICD-9-CM | 238.3 | Neoplasm of uncertain behavior of breast |
| ICD-9-CM | 239.3 | Neoplasm of unspecified nature of breast |
| ICD-9-CM | V10.3 | Personal history of malignant neoplasm of breast |
| Breast cancer sign or symptom codes | | |
| Type | Code | Descriptor |
| ICD-9-CM | 611.72 | Lump or mass in breast |
| ICD-10-CM | N63.0 | Unspecified lump in unspecified breast |
| ICD-10-CM | N63.10 | Unspecified lump in the right breast, unspecified quadrant |
| ICD-10-CM | N63.11 | Unspecified lump in the right breast, upper outer quadrant |
| ICD-10-CM | N63.12 | Unspecified lump in the right breast, upper inner quadrant |
| ICD-10-CM | N63.13 | Unspecified lump in the right breast, lower outer quadrant |
| ICD-10-CM | N63.14 | Unspecified lump in the right breast, lower inner quadrant |
| ICD-10-CM | N63.15 | Unspecified lump in the right breast, overlapping quadrants |
| ICD-10-CM | N63.20 | Unspecified lump in the left breast, unspecified quadrant |
| ICD-10-CM | N63.21 | Unspecified lump in the left breast, upper outer quadrant |
| ICD-10-CM | N63.22 | Unspecified lump in the left breast, upper inner quadrant |
| ICD-10-CM | N63.23 | Unspecified lump in the left breast, lower outer quadrant |
| ICD-10-CM | N63.24 | Unspecified lump in the left breast, lower inner quadrant |
| ICD-10-CM | N63.25 | Unspecified lump in the left breast, overlapping quadrants |
| ICD-10-CM | N63.31 | Unspecified lump in axillary tail of the right breast |
| ICD-10-CM | N63.32 | Unspecified lump in axillary tail of the left breast |
| ICD-10-CM | N63.41 | Unspecified lump in right breast, subareolar |
| ICD-10-CM | N63.42 | Unspecified lump in left breast, subareolar |
| ICD-9-CM | 611.71 | Mastodynia |
| ICD-10-CM | N64.4 | Mastodynia |
| ICD-9-CM | 611.79 | Other signs and symptoms in breast |
| ICD-10-CM | N64.51 | Induration of breast |
| ICD-10-CM | N64.52 | Nipple discharge |
| ICD-10-CM | N64.53 | Retraction of nipple |

| | | |
|---|---|---|
| ICD-10-CM | N64.59 | Other signs and symptoms in breast |
| ICD-9-CM | 757.6 | Specified congenital anomalies of breast |
| ICD-10-CM | Q83.0 | Congenital absence of breast with absent nipple |
| ICD-10-CM | Q83.1 | Accessory breast |
| ICD-10-CM | Q83.2 | Absent nipple |
| ICD-10-CM | Q83.3 | Accessory nipple |
| ICD-10-CM | Q83.8 | Other congenital malformations of breast |
| ICD-10-CM | Q83.9 | Congenital malformation of breast, unspecified |
| Breast involvement codes | | |
| Type | Code | Descriptor |
| ICD-9-CM | 196.3 | Secondary and unspecified malignant neoplasm of lymph nodes of axilla and upper limb |
| ICD-10-CM | C77.3 | Secondary and unspecified malignant neoplasm of axilla and upper limb lymph nodes |
| ICD-9-CM | 217 | Benign neoplasm of breast |
| ICD-10-CM | D24.1 | Benign neoplasm of right breast |
| ICD-10-CM | D24.2 | Benign neoplasm of left breast |
| ICD-10-CM | D24.9 | Benign neoplasm of unspecified breast |
| ICD-9-CM | 610.0 | Solitary cyst of breast |
| ICD-9-CM | 610.1 | Diffuse cystic mastopathy |
| ICD-9-CM | 610.2 | Fibroadenosis of breast |
| ICD-9-CM | 610.3 | Fibrosclerosis of breast |
| ICD-9-CM | 610.4 | Mammary duct ectasia |
| ICD-9-CM | 610.8 | Other specified benign mammary dysplasias |
| ICD-9-CM | 610.9 | Benign mammary dysplasia, unspecified |
| ICD-10-CM | N60.01 | Solitary cyst of right breast |
| ICD-10-CM | N60.02 | Solitary cyst of left breast |
| ICD-10-CM | N60.09 | Solitary cyst of unspecified breast |
| ICD-10-CM | N60.11 | Diffuse cystic mastopathy of right breast |
| ICD-10-CM | N60.12 | Diffuse cystic mastopathy of left breast |
| ICD-10-CM | N60.19 | Diffuse cystic mastopathy of unspecified breast |
| ICD-10-CM | N60.21 | Fibroadenosis of right breast |
| ICD-10-CM | N60.22 | Fibroadenosis of left breast |
| ICD-10-CM | N60.29 | Fibroadenosis of unspecified breast |
| ICD-10-CM | N60.31 | Fibrosclerosis of right breast |
| ICD-10-CM | N60.32 | Fibrosclerosis of left breast |
| ICD-10-CM | N60.39 | Fibrosclerosis of unspecified breast |
| ICD-10-CM | N60.41 | Mammary duct ectasia of right breast |
| ICD-10-CM | N60.42 | Mammary duct ectasia of left breast |
| ICD-10-CM | N60.49 | Mammary duct ectasia of unspecified breast |
| ICD-10-CM | N60.81 | Other benign mammary dysplasias of right breast |
| ICD-10-CM | N60.82 | Other benign mammary dysplasias of left breast |
| ICD-10-CM | N60.89 | Other benign mammary dysplasias of unspecified breast |
| ICD-10-CM | N60.91 | Unspecified benign mammary dysplasia of right breast |

| | | |
|---|---|---|
| ICD-10-CM | N60.92 | Unspecified benign mammary dysplasia of left breast |
| ICD-10-CM | N60.99 | Unspecified benign mammary dysplasia of unspecified breast |
| ICD-9-CM | 611.0 | Inflammatory disease of breast |
| ICD-10-CM | N61.0 | Mastitis without abscess |
| ICD-10-CM | N61.1 | Abscess of the breast and nipple |
| ICD-10-CM | N61.20 | Granulomatous mastitis, unspecified breast |
| ICD-10-CM | N61.21 | Granulomatous mastitis, right breast |
| ICD-10-CM | N61.22 | Granulomatous mastitis, left breast |
| ICD-10-CM | N61.23 | Granulomatous mastitis, bilateral breast |
| ICD-9-CM | 611.1 | Hypertrophy of breast |
| ICD-10-CM | N62 | Hypertrophy of breast |
| ICD-9-CM | 611.2 | Fissure of nipple |
| ICD-10-CM | N64.0 | Fissure and fistula of nipple |
| ICD-9-CM | 611.3 | Fat necrosis of breast |
| ICD-10-CM | N64.1 | Fat necrosis of breast |
| ICD-9-CM | 611.4 | Atrophy of breast |
| ICD-10-CM | N64.2 | Atrophy of breast |
| ICD-9-CM | 611.5 | Galactocele |
| ICD-10-CM | N64.89 | Other specified disorders of breast |
| ICD-9-CM | 611.6 | Galactorrhea not associated with childbirth |
| ICD-10-CM | N64.3 | Galactorrhea not associated with childbirth |
| ICD-9-CM | 611.81 | Ptosis of breast |
| ICD-9-CM | 611.82 | Hypoplasia of breast |
| ICD-9-CM | 611.83 | Capsular contracture of breast implant |
| ICD-9-CM | 611.89 | Other specified disorders of breast |
| ICD-10-CM | N64.81 | Ptosis of breast |
| ICD-10-CM | N64.82 | Hypoplasia of breast |
| ICD-10-CM | N64.89 | Other specified disorders of breast |
| ICD-9-CM | 611.9 | Unspecified breast disorder |
| ICD-10-CM | N64.9 | Disorder of breast, unspecified |
| ICD-9-CM | 793.80 | Abnormal mammogram, unspecified |
| ICD-9-CM | 793.81 | Mammographic microcalcification |
| ICD-9-CM | 793.82 | Inconclusive mammogram |
| ICD-9-CM | 793.89 | Other (abnormal) findings on radiological examination of breast |
| ICD-10-CM | R92.8 | Other abnormal and inconclusive findings on diagnostic imaging of breast |
| ICD-9-CM | V16.3 | Family history of malignant neoplasm of breast |
| ICD-10-CM | Z80.3 | Family history of malignant neoplasm of breast |
| Bilateral mastectomy codes | | |
| Type | Code | Description |
| ICD-10-PCS | 0HTV0ZZ | Resection of Bilateral Breast, Open Approach |
| ICD-9-PCS | 85.42 | Bilateral simple mastectomy |
| ICD-9-PCS | 85.44 | Bilateral extended simple mastectomy |
| ICD-9-PCS | 85.46 | Bilateral radical mastectomy |

| | | |
|---|---|---|
| ICD-9-PCS | 85.48 | Bilateral extended radical mastectomy |
| ICD-10-CM | Z90.13 | Acquired absence of bilateral breasts and nipples |
| ICD-9-CM | V45.71 | Acquired absence of breast and nipple |
| CPT | 19303† | Mastectomy, simple, complete |
| CPT | 19304† | Mastectomy, subcutaneous |
| CPT | 19305† | Mastectomy, radical, including pectoral muscles, axillary lymph nodes |
| CPT | 19306† | Mastectomy, radical, including pectoral muscles, axillary and internal mammary lymph nodes (Urban type operation) |
| CPT | 19307† | Mastectomy, radical, including pectoral muscles, axillary and internal mammary lymph nodes (Urban type operation) |
| Unilateral mastectomy codes with bilateral modifiers | | |
| Type | Code | Description |
| ICD-10-PCS | 0HTU0ZZ* | Resection of Left Breast, Open Approach |
| ICD-10-PCS | 0HTT0ZZ** | Resection of Right Breast, Open Approach |
| ICD-9-PCS | 85.41‡ | Unilateral simple mastectomy |
| ICD-9-PCS | 85.43‡ | Unilateral extended simple mastectomy |
| ICD-9-PCS | 85.45‡ | Unilateral radical mastectomy |
| ICD-9-PCS | 85.47‡ | Unilateral extended radical mastectomy |
| ICD-10-CM | Z90.12* | Acquired absence of left breast and nipple |
| ICD-10-CM | Z90.11** | Acquired absence of right breast and nipple |
| CPT | 19303*** | Mastectomy, simple, complete |
| CPT | 19304*** | Mastectomy, subcutaneous |
| CPT | 19305*** | Mastectomy, radical, including pectoral muscles, axillary lymph nodes |
| CPT | 19306*** | Mastectomy, radical, including pectoral muscles, axillary and internal mammary lymph nodes (Urban type operation) |
| CPT | 19307*** | Mastectomy, radical, including pectoral muscles, axillary and internal mammary lymph nodes (Urban type operation) |

† With bilateral modifier -50 on the same procedure
*With right-side modifier -RT and code on same day or different service days
**With left-side modifier -LT and code on same day or different service days
***With left-side modifier -LT and right-side modifier -RT on same day or different service days
‡ Two separate occurrences on two different dates of service
Note: Only CPT and HCPCS codes effective during the study period (Jan 1, 2010 thru Dec 31, 2019) were included for analysis. Other codes for screening (e.g., 19180,19200,19220,19240) were not included in the analysis.

Source:(1-5, 7-9)

### Colorectal Cancer Screening

#### *Colorectal Cancer Screening Inclusion Codes*

*Table S10 List of Procedure and Diagnosis codes for Colorectal Cancer Screening*

| Flexible Sigmoidoscopy Codes | | |
|---|---|---|
| Type | CPT | Description |
| CPT | 45330 | Sigmoidoscopy, flexible; diagnostic, including collection of specimen(s) by brushing or washing, when performed (separate procedure) |
| CPT | 45331 | Sigmoidoscopy, flexible with biopsy, single or multiple |
| CPT | 45332 | Sigmoidoscopy, flexible with removal of foreign body(s) |
| CPT | 45333 | Sigmoidoscopy, flexible with removal of tumor(s), polyp(s), or other lesion(s) by hot biopsy forceps |
| CPT | 45334 | Sigmoidoscopy, flexible; with control of bleeding, any method |
| CPT | 45335 | Sigmoidoscopy, flexible; with directed submucosal injection(s), any substance |
| CPT | 45337 | Sigmoidoscopy, flexible; with decompression (for pathologic distention) (eg, volvulus, megacolon), including placement of decompression tube, when performed |
| CPT | 45338 | Sigmoidoscopy, flexible with removal of tumor(s), polyp(s), or other lesion(s) by snare technique |
| CPT | 45339 | Sigmoidoscopy, flexible; with ablation of tumor(s), polyp(s), or other lesion(s) not amenable to removal by hot biopsy forceps, bipolar cautery or snare technique |
| CPT | 45340 | Sigmoidoscopy, flexible with transendoscopic balloon dilation |
| CPT | 45341 | Sigmoidoscopy, flexible with endoscopic ultrasound examination |
| CPT | 45342 | Sigmoidoscopy, flexible; with transendoscopic ultrasound guided intramural or transmural fine needle aspiration/biopsy(s) |
| CPT | 45345 | Sigmoidoscopy, flexible; with transendoscopic stent placement (includes predilation) |
| CPT | 45346 | Sigmoidoscopy, flexible; with ablation of tumor(s), polyp(s), or other lesion(s) (includes pre- and post-dilation and guide wire passage, when performed) |
| CPT | 45347 | Sigmoidoscopy, flexible; with placement of endoscopic stent (includes pre- and post-dilation and guide wire passage, when performed) |
| CPT | 45349 | Sigmoidoscopy, flexible with endoscopic mucosal resection |
| CPT | 45350 | Sigmoidoscopy, flexible with band ligation(s) (eg, hemorrhoids) |
| CPT | 45300 | Proctosigmoidoscopy, rigid; diagnostic, with or without collection of specimen(s) by brushing or washing (separate procedure) |
| CPT | 45305 | Proctosigmoidoscopy, rigid; with biopsy, single or multiple |
| CPT | 45308 | Proctosigmoidoscopy, rigid; with removal of single tumor, polyp, or other lesion by hot biopsy forceps or bipolar cautery |

| | | |
|---|---|---|
| CPT | 45309 | Proctosigmoidoscopy, rigid; with removal of single tumor, polyp, or other lesion by snare technique |
| CPT | 45315 | Proctosigmoidoscopy, rigid; with removal of multiple tumors, polyps, or other lesions by hot biopsy forceps, bipolar cautery or snare technique |
| CPT | 45320 | Proctosigmoidoscopy, rigid; with ablation of tumor(s), polyp(s), or other lesion(s) not amenable to removal by hot biopsy forceps, bipolar cautery or snare technique (eg, laser) |
| HCPCS | S0601 | Screening proctoscopy |
| HCPCS | G0104 | Colorectal cancer screening flexible sigmoidoscopy |
| HCPCS | G6022 | Sigmoidoscopy, flexible; with ablation of tumor(s), polyp(s), or other lesions(s) not amenable to removal by hot biopsy forceps, bipolar cautery or snare technique |
| ICD-9-PCS | 45.24 | Flexible sigmoidoscopy |
| Colonoscopy codes | | |
| CPT | 44388 | Colonoscopy through stoma; diagnostic, including collection of specimen(s) by brushing or washing, when performed (separate procedure) |
| CPT | 44389 | Colonoscopy through stoma with biopsy, single or multiple |
| CPT | 44390 | Colonoscopy through stoma with removal of foreign body(s) |
| CPT | 44391 | Colonoscopy through stoma with control of bleeding, any method |
| CPT | 44392 | Colonoscopy through stoma with removal of tumor(s), polyp(s), or other lesion(s) by hot biopsy forceps |
| CPT | 44393 | Colonoscopy through stoma; with ablation of tumor(s), polyp(s), or other lesion(s) not amenable to removal by hot biopsy forceps, bipolar cautery or snare technique |
| CPT | 44394 | Colonoscopy through stoma with removal of tumor(s), polyp(s), or other lesion(s) by snare technique |
| CPT | 44397 | Colonoscopy through stoma; with transendoscopic stent placement (includes predilation) |
| CPT | 44401 | Colonoscopy through stoma; with ablation of tumor(s), polyp(s), or other lesion(s) (includes pre-and post-dilation and guide wire passage, when performed) |
| CPT | 44402 | Colonoscopy through stoma with endoscopic stent placement (including pre- and post-dilation and guide wire passage, whe... |
| CPT | 44403 | Colonoscopy through stoma with endoscopic mucosal resection |
| CPT | 44404 | Colonoscopy through stoma with directed submucosal injection(s), any substance |
| CPT | 44405 | Colonoscopy through stoma with transendoscopic balloon dilation |
| CPT | 44406 | Colonoscopy through stoma with endoscopic ultrasound examination, limited to the sigmoid, descending, transverse, or as... |
| CPT | 44407 | Colonoscopy through stoma with transendoscopic ultrasound guided intramural or transmural fine needle aspiration/biopsy... |

| | | |
|---|---|---|
| CPT | 44408 | Colonoscopy through stoma with decompression (for pathologic distention) (eg, volvulus, megacolon), including placement... |
| CPT | 45355 | Colonoscopy, rigid or flexible, transabdominal via colotomy, single or multiple |
| CPT | 45378 | Colonoscopy, flexible; diagnostic, including collection of specimen(s) by brushing or washing, when performed (separate procedure) |
| CPT | 45379 | Colonoscopy, flexible with removal of foreign body(s) |
| CPT | 45380 | Colonoscopy, flexible with biopsy, single or multiple |
| CPT | 45381 | Colonoscopy, flexible with directed submucosal injection(s), any substance |
| CPT | 45382 | Colonoscopy, flexible with control of bleeding, any method |
| CPT | 45383 | Colonoscopy, flexible, proximal to splenic flexure; with ablation of tumor(s), polyp(s), or other lesion(s) not amenable to removal by hot biopsy forceps, bipolar cautery or snare technique |
| CPT | 45384 | Colonoscopy, flexible with removal of tumor(s), polyp(s), or other lesion(s) by hot biopsy forceps |
| CPT | 45385 | Colonoscopy, flexible with removal of tumor(s), polyp(s), or other lesion(s) by snare technique |
| CPT | 45386 | Colonoscopy, flexible with transendoscopic balloon dilation |
| CPT | 45387 | Colonoscopy, flexible, proximal to splenic flexure; with transendoscopic stent placement (includes predilation) |
| CPT | 45388 | Colonoscopy, flexible; with ablation of tumor(s), polyp(s), or other lesion(s) (includes pre- and post-dilation and guide wire passage, when performed) |
| CPT | 45389 | Colonoscopy, flexible with endoscopic stent placement (includes pre- and post-dilation and guide wire passage, when per... |
| CPT | 45390 | Colonoscopy, flexible with endoscopic mucosal resection |
| CPT | 45391 | Colonoscopy, flexible; with endoscopic ultrasound examination limited to the rectum, sigmoid, descending, transverse, or ascending colon and cecum, and adjacent structures |
| CPT | 45392 | Colonoscopy, flexible; with transendoscopic ultrasound guided intramural or transmural fine needle aspiration/biopsy(s), includes endoscopic ultrasound examination limited to the rectum, sigmoid, descending, transverse, or ascending colon and cecum, and adjacent structures |
| CPT | 45393 | Colonoscopy, flexible; with decompression (for pathologic distention) (eg, volvulus, megacolon), including placement of decompression tube, when performed |
| CPT | 45398 | Colonoscopy, flexible with band ligation(s) (eg, hemorrhoids) |
| HCPCS | G0105 | Colorectal cancer screening colonoscopy on individual at high risk |
| HCPCS | G0121 | Colorectal cancer screening colonoscopy on individual not meeting criteria for high risk |
| HCPCS | G6024 | Colonoscopy, flexible; proximal to splenic flexure; with ablation of tumor(s), polyp(s), or other lesion(s) not amenable to removal by hot biopsy forceps, bipolar cautery or snare tehnique |

| | | |
|---|---|---|
| ICD-9-PCS | 45.22 | Endoscopy of large intestine through artificial stoma |
| ICD-9-PCS | 45.23 | Colonoscopy |
| ICD-9-PCS | 45.25 | Closed [endoscopic] biopsy of large intestine |
| ICD-9-PCS | 45.42 | Endoscopic polypectomy of large intestine |
| ICD-9-PCS | 45.43 | Endoscopic destruction of other lesion or tissue of large intestine |
| ICD-10-PCS | 0DJD8ZZ | Inspection of Lower Intestinal Tract, Via Natural or Artificial Opening Endoscopic |
| Computed tomographic (CT) Colonography codes | | |
| CPT | 74261 | Computed tomographic (CT) colonography, diagnostic, including image postprocessing without contrast material |
| CPT | 74262 | Computed tomographic (CT) colonography, diagnostic, including image postprocessing; with contrast material(s) including non-contrast images, if performed |
| CPT | 74263 | Computed tomographic (CT) colonography, screening, including image postprocessing |
| HCPCS | 0066T | Computed tomographic [CT] colonography [i.e., virtual colonoscopy]; screening |
| HCPCS | 0067T | Computed Tomographic Colonography (virtual colonoscopy) diagnostic |
| Guaiac fecal occult blood test (gFOBT) codes | | |
| CPT | 82270 | Blood, occult, by peroxidase activity (eg, guaiac), qualitative; feces, consecutive collected specimens with single determination, for colorectal neoplasm screening (ie, patient was provided 3 cards or single triple card for consecutive collection) |
| HCPCS | G0328 | Colorectal cancer screening fecal occult blood test, immunoassay, 1-3 simultaneous |
| Fecal immunochemical test (FIT) codes | | |
| CPT | 82274 | Blood, occult, by fecal hemoglobin determination by immunoassay, qualitative, feces, 1-3 simultaneous determinations |
| Double contrast barium enema (DCBE) codes | | |
| HCPCS | G0106 | Colorectal cancer screening; alternative to G0104, screening sigmoidoscopy, barium enema |
| HCPCS | G0122 | Colorectal cancer screening; barium enema |
| CPT | 74270 | Radiologic examination, colon, including scout abdominal radiograph(s) and delayed image(s), when performed; single-contrast (eg, barium) study |
| CPT | 74280 | Radiologic examination, colon, including scout abdominal radiograph(s) and delayed image(s), when performed; double-contrast (eg, high density barium and air) study, including glucagon, when administered |

| | | |
|---|---|---|
| HCPCS | G0120 | Colorectal cancer screening alternative to G0105, screening colonoscopy, barium enema. |
| sDNA-FIT (Cologuard) codes | | |
| HCPCS | 81528 | Oncology (colorectal) screening, quantitative real-time target and signal amplification of 10 DNA markers (KRAS mutations, promoter methylation of NDRG4 and BMP3) and fecal hemoglobin, utilizing stool, algorithm reported as a positive or negative result |
| HCPCS | G0464 | Colorectal cancer screening; stool-based dna and fecal occult hemoglobin (e.g., kras, ndrg4 and bmp3 |
| HCPCS | S3890 | Dna analysis, fecal, for colorectal cancer screening |

sDNA-FIT: multitarget stool DNA with FIT component; FIT: fecal immunochemical test; gFOBT: Guaiac fecal occult blood test; CT: Computed tomographic; DCBE: Double contrast barium enema

Source: (2, 8-13)

*Colorectal Cancer Screening Exclusion Codes*

*Table S11 List of Exclusionary Procedure and Diagnosis Codes for Colorectal Cancer, including Total Colectomy and Evidence of Prior Colorectal Cancer Diagnosis*

| Colorectal Cancer | | |
|---|---|---|
| Type | Code | Descriptor |
| ICD-10-CM | C18.0 | Malignant neoplasm of cecum |
| ICD-10-CM | C18.1 | Malignant neoplasm of appendix |
| ICD-10-CM | C18.2 | Malignant neoplasm of ascending colon |
| ICD-10-CM | C18.3 | Malignant neoplasm of hepatic flexure |
| ICD-10-CM | C18.4 | Malignant neoplasm of transverse colon |
| ICD-10-CM | C18.5 | Malignant neoplasm of splenic flexure |
| ICD-10-CM | C18.6 | Malignant neoplasm of descending colon |
| ICD-10-CM | C18.7 | Malignant neoplasm of sigmoid colon |
| ICD-10-CM | C18.8 | Malignant neoplasm of overlapping sites of colon |
| ICD-10-CM | C18.9 | Malignant neoplasm of colon, unspecified |
| ICD-10-CM | C19 | Malignant neoplasm of rectosigmoid junction |
| ICD-10-CM | C20 | Malignant neoplasm of rectum |
| ICD-10-CM | C21.2 | Malignant neoplasm of cloacogenic zone |
| ICD-10-CM | C21.8 | Malignant neoplasm of overlapping sites of rectum, anus and anal canal |
| ICD-10-CM | C78.5 | Secondary malignant neoplasm of large intestine and rectum |
| ICD-10-CM | Z85.038 | Personal history of other malignant neoplasm of large intestine |
| ICD-10-CM | Z85.048 | Personal history of other malignant neoplasm of rectum, rectosigmoid junction, and anus |
| ICD-9-CM | 153.0 | Malignant neoplasm of hepatic flexure |
| ICD-9-CM | 153.1 | Malignant neoplasm of transverse colon |
| ICD-9-CM | 153.2 | Malignant neoplasm of descending colon |
| ICD-9-CM | 153.3 | Malignant neoplasm of sigmoid colon |
| ICD-9-CM | 153.4 | Malignant neoplasm of cecum |
| ICD-9-CM | 153.5 | Malignant neoplasm of appendix vermiformis |
| ICD-9-CM | 153.6 | Malignant neoplasm of ascending colon |
| ICD-9-CM | 153.7 | Malignant neoplasm of splenic flexure |
| ICD-9-CM | 153.8 | Malignant neoplasm of other specified sites of large intestine |
| ICD-9-CM | 153.9 | Malignant neoplasm of colon, unspecified site |
| ICD-9-CM | 154.0 | Malignant neoplasm of rectosigmoid junction |
| ICD-9-CM | 154.1 | Malignant neoplasm of rectum |
| ICD-9-CM | 154.2 | Malignant neoplasm of anal canal |
| ICD-9-CM | 154.3 | Malignant neoplasm of anus, unspecified site |
| ICD-9-CM | 197.5 | Secondary malignant neoplasm of large intestine and rectum |

| | | |
|---|---|---|
| ICD-9-CM | V10.05 | Personal history of malignant neoplasm of large intestine |
| ICD-9-CM | V10.06 | Personal history of malignant neoplasm of rectum, rectosigmoid junction, and anus |
| Total Colectomy | | |
| Type | Code | Descriptor |
| CPT | 44150 | Colectomy, total, abdominal, without proctectomy; with ileostomy or ileoproctostomy |
| CPT | 44151 | Colectomy, total, abdominal, without proctectomy; with continent ileostomy |
| CPT | 44152 | Colectomy, total, abdominal, without proctectomy; with rectal mucosectomy, ileoanal anastomosis, with or without loop ileostomy. |
| CPT | 44153 | Colectomy, total, abdominal, without proctectomy; with rectal mucosectomy, ileoanal anastomosis, creation of ileal reservoir (S or J), with or without loop ileostomy. |
| CPT | 44155 | Colectomy, total, abdominal, with proctectomy; with ileostomy |
| CPT | 44156 | Colectomy, total, abdominal, with proctectomy; with continent ileostomy |
| CPT | 44157 | Colectomy, total, abdominal, with proctectomy; with ileoanal anastomosis, includes loop ileostomy, and rectal mucosectomy, when performed |
| CPT | 44158 | Colectomy, total, abdominal, with proctectomy; with ileoanal anastomosis, creation of ileal reservoir (S or J), includes loop ileostomy, and rectal mucosectomy, when performed |
| CPT | 44210 | Laparoscopy, surgical; colectomy, total, abdominal, without proctectomy, with ileostomy or ileoproctostomy |
| CPT | 44211 | Laparoscopy, surgical; colectomy, total, abdominal, with proctectomy, with ileoanal anastomosis, creation of ileal reservoir (S or J), with loop ileostomy, includes rectal mucosectomy, when performed |
| CPT | 44212 | Laparoscopy, surgical; colectomy, total, abdominal, with proctectomy, with ileostomy |
| ICD-10-PCS | 0DTE0ZZ | Resection of Large Intestine, Open Approach |
| ICD-10-PCS | 0DTE4ZZ | Resection of Large Intestine, Percutaneous Endoscopic Approach |
| ICD-10-PCS | 0DTE7ZZ | Resection of Large Intestine, Via Natural or Artificial Opening |
| ICD-10-PCS | 0DTE8ZZ | Resection of Large Intestine, Via Natural or Artificial Opening Endoscopic |
| ICD-9-PCS | 45.81 | Laparoscopic total intra-abdominal colectomy |
| ICD-9-PCS | 45.82 | Open total intra-abdominal colectomy |
| ICD-9-PCS | 45.83 | Other and unspecified total intra-abdominal colectomy |
| HCPCS | G9711 | Patients with a diagnosis or past history of total colectomy or colorectal cancer |
| HCPCS | G0213 | Pet imaging whole body; diagnosis; colorectal [G0213] |
| HCPCS | G0214 | Pet imaging whole body; initial staging; colorectal [G0214] |

| | | |
|---|---|---|
| HCPCS | G0215 | Pet imaging whole body; restaging; colorectal cancer [G0215] |
| HCPCS | G0231 | Pet, whole body, for recurrence of colorectal or colorectal metastatic cancer; gamma cameras only [G0231] |

Sources: (2, 8-13)
